# COVID-19: One-month impact of the French lockdown on the epidemic burden

**DOI:** 10.1101/2020.04.22.20075705

**Authors:** Jonathan Roux, Clément Massonnaud, Pascal Crépey

## Abstract

On March 16 2020, French authorities ordered a large scale lockdown to counter the COVID-19 epidemic wave rising in the country, stopping non-essential economic, educational, and entertainment activities, maintaining mainly food retailers and healthcare institutions. One month later, the number of new hospitalizations and ICU admissions had reached a plateau and were beginning a slow descent.

We developed a spatialized, deterministic, age-structured, and compartmental SARS-CoV-2 transmission model able to reproduce the pre-lockdown dynamic of the epidemic in each of the 13 French metropolitan regions. Thanks to this model, we estimate, at regional and national levels, the total number of hospitalizations, ICU admissions, hospital beds requirements (hospitalization and ICU), and hospital deaths which may have been prevented by this massive and unprecedented intervention in France.

If no control measures had been set up, between March 19 and April 19 2020, our analysis shows that almost 23% of the French population would have been affected by COVID-19 (14.8 million individuals). Hence, the French lockdown prevented 587,730 hospitalizations and 140,320 ICU admissions at the national level. The total number of ICU beds required to treat patients in critical conditions would have been 104,550, far higher than the maximum French ICU capacity. This first month of lockdown also permitted to avoid 61,739 hospital deaths, corresponding to a 83.5% reduction of the total number of predicted deaths.

Our analysis shows that in absence of any control measures, the COVID-19 epidemic would have had a critical morbidity and mortality burden in France, overwhelming in a matter of weeks French hospital capacities.

## 2 Introduction

The World Health Organization (WHO) published its first alert on the novel coronavirus SARS-CoV-2 in early January,^1^ and declared the COVID-19 situation was a pandemic on March 11, 2020.^2^ In the meantime, travel restrictions and containment strategies had failed to prevent SARS-CoV-2 from becoming a global threat, and COVID-19 cases and deaths were reported in most countries.

In France, the first cases of COVID-19 were confirmed on January 24.^3^ A first cluster of cases was detected on February 8 in Haute Savoie, later followed by a second one in Oise, and a third in Brittany. France became one of the most impacted country in Europe, behind Italy and Spain. On March 12, the closure of schools and universities had to be ordered, followed, four days later, by the announcement of a full lockdown of the French population starting on March 17.^4^

Several models were developed in an attempt to forecast the transmission dynamic of the virus in France. Although these models were initially simplistic, and relied on the limited data available at the time, these preliminary analyses were helpful in predicting various epidemiological outcomes. Our first work sought to use a deterministic compartmental model to forecast hospital needs for each of the 13 French metropolitan regions.^5^ Like others, our model predicted that, without strong control measures, the epidemic would overwhelm the French healthcare system, much like it was overwhelming the Italian one.

Lockdown measures proved to be effective in controlling the outbreak in China.^6,7^ However, whether they would have a similar impact in European countries remained unclear. Several studies tried to estimate the potential impact of various control measures in European countries. Di Domenico and colleagues proposed a stochastic model to forecast the expected impact of school closures and telework measure in three French regions.^8^ Ferguson and colleagues used an individual based simulation model to assess the potential effectiveness of mitigation and suppression strategies in Great Britain and the United States.^9^ Both predicted that mild control strategies would be insufficient to successfully mitigate the epidemic. But the expected impact of control measures such as a lockdown, was necessarily subject to a lot of uncertainty, as it greatly depends on the level of reduction of contacts that could be achieved, which is hard to estimate.

Lockdown measures, which have been set up in most European countries, triggered massive distraught on populations and economies. Evaluating the impact of these strategies on the different epidemiological outcomes is therefore of the utmost importance. As more data is gathered, we are able to provide more detailed epidemiological description of the epidemic, and better insights about its dynamic. This new context allows us to refine preliminary forecasts, by using more sophisticated models.

This analysis attempts to retrospectively estimate the effect of a one-month long lockdown in France on hospital requirements and mortality, using a compartmental, deterministic mathematical model.

## 3 Methods

### 3.1 Epidemiological data for France

We retrieved epidemiological regional data related to the COVID-19 epidemic in metropolitan France gathered by the French National Public Health Agency (SpF-’Santé publique France’) and freely available online.^10^ On the one side, we collected incidence data on the daily number of hospital admissions (non-ICU and ICU considered separately), and the daily number of deaths in hospitals (deaths in nursing homes and at home were not considered). On the other side, we also compiled prevalence data on the daily number of occupied beds (non-ICU and ICU). In addition, we obtained from the French Directorate for Research, Studies, Evaluation and Statistics (Drees-’Direction de la recherche, des études, de l’évaluation et des statistiques’) the daily number of patient transfers between regions (personal communication) in order to adjust the forecasts of the model to real settings.

The COVID-19 epidemic did not have a synchronous dynamic among the different French regions, mainly due to different timings of introduction, super-spreading events, or proximity with countries where the virus was already circulating. Hence, we divided metropolitan France into its 13 metropolitan Regions [French administrative areas with an averaged population of 4.75 millions ranging from 300,000 (Corse) to 12.55 millions (Ile-de-France)]. For each Region, we computed the corresponding catchment area using Voronoi polygons (Figure S1). Population structure was inferred for each catchment area from 2016 and 2017 census data provided by the French National Institute of Statistics and Economic Studies (Insee).^11,12^

Data on ICU beds capacity per French Region were retrieved from the Drees regarding the current situation on April 2, 2020 (personal communication) and from the annual statistical survey of French healthcare facilities (SAE-’Statistique Annuelle des Etablissements’) regarding the baseline capacity.

### 3.2 SARS-CoV-2 transmission model

We developed a deterministic, age-structured, compartmental model (Figure 1). The population was divided into 17 age-groups: 16 age-band of 5 years from 0 to 80 years, and a last group for people aged 80 years and older. Initially, we consider that individuals are susceptible (*S*), and then potentially exposed to the virus but not infectious (*E*). Exposed individuals may become infectious while being in a pre-symptomatic stage (*I*_*ps*_), or remain asymptomatic (*As*). Pre-symptomatic individuals will then become infected symptomatic (*I*_*s*_) before being either hospitalized (*I*_*h*_) or remain within the community (*I*_*nh*_). Patients cured or dying from COVID-19 are finally removed from the chain of infection (*R*).

**Figure 1:**
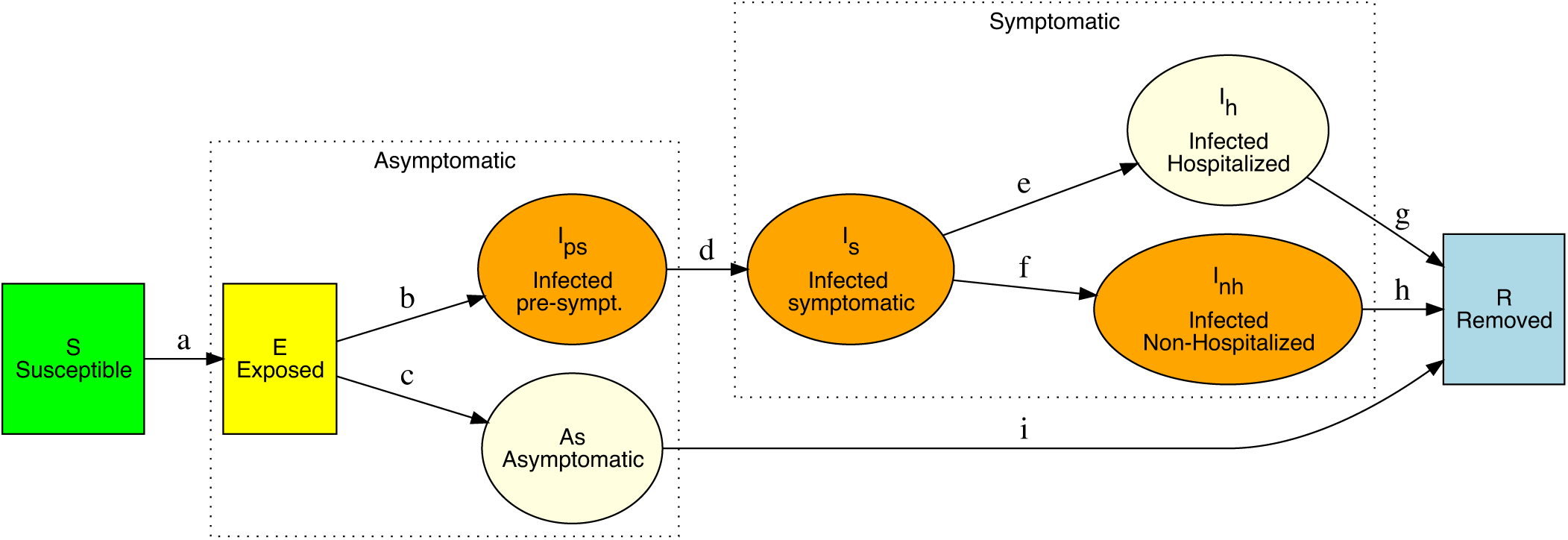
Diagram of the SARS-CoV-2 epidemiological model.

Transitions between compartments are labeled from *a* to *e* and described in Table 1.

**Table 1:**
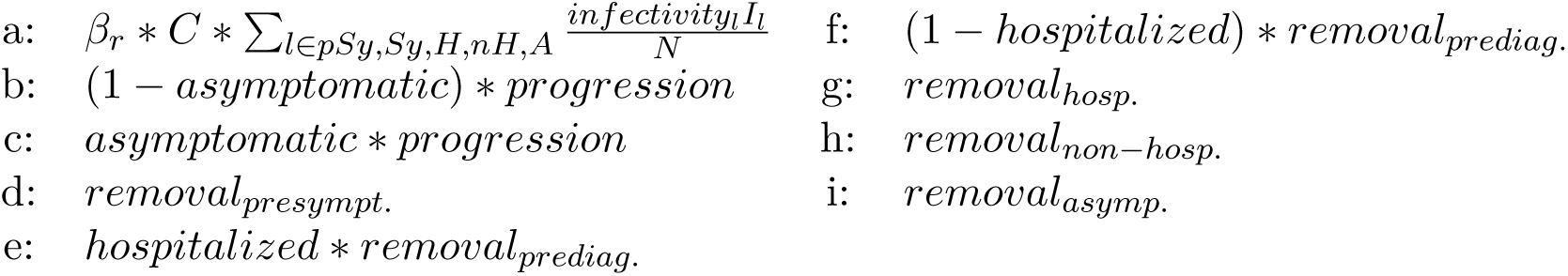
Definitions of transitions between compartments of the SARS-CoV-2 transmission model. *C* stands for the inter-individual contact matrix, and *β*_*r*_ for the probability of infection upon contact for each Region *r*.

### 3.3 Epidemiological parameters

#### 3.3.1 Transmission model parameters

The parameters used in our transmission model were selected from the available literature on the COVID-19 pandemic and are gathered in the Table 2. We set the incubation period to 5.1 days,^9^ and deduced the duration of the pre-symptomatic infectious phase (2.38 days) from the pre-symptomatic incubation period of 2.72 days.^13^ We fixed the length of the symptomatic phase to 8 days and assumed the pre-diagnostic phase was equal to 1.5 days, leading to a diagnosed symptomatic phase of 6.5 days. Finally, we deduced the average duration of the asymptomatic phase by summing the pre-symptomatic incubation period and the symptomatic phase altogether, namely 10.72 days.

**Table 2:**
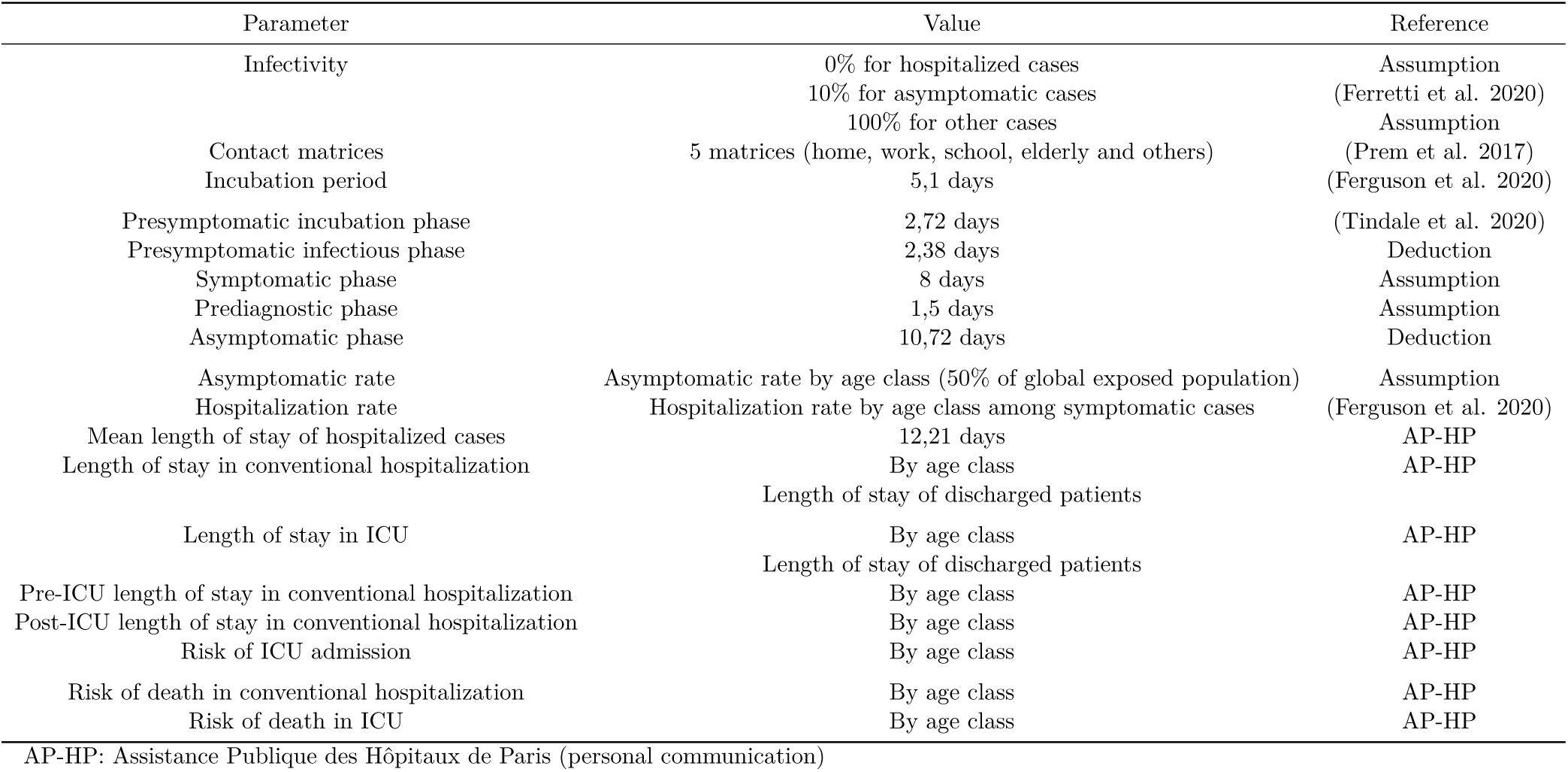
Table of epidemiological parameters used in the model.

To compute the number of infected cases needing hospitalization, we used the rates of hospitalization of symptomatic cases suggested by Ferguson et al..^9^ An age-dependent risk of being asymptomatic was estimated using the age-dependent complementary risks of being a severe case when infected proposed by Guan et al.,^14^ which were multiplied by an estimated coefficient to reach a global proportion of 50% of asymptomatic in the population of exposed cases.

Regarding the infectivity of cases in each compartment, based on the work of Ferretti et al.,^15^ we fixed that asymptomatic cases were 90% less infectious than symptomatic ones. We also assumed that infected hospitalized cases were no longer able to infect other individuals due to their isolation in hospital rooms and the protection of the hospital’s staff. We acknowledge that this last assumption is in contradiction with the observed rate of infection among healthcare workers, but we assume that their infection will have no significant impact on the overall epidemic dynamic. All other compartments were considered as having a 100% infectivity rate.

#### 3.3.2 Contact matrices

To simulate age and location-dependent mixing, we used inter-individual contacts matrices for the French population estimated by Prem et al. for contacts at home, work, school and in other locations.^16^ We isolated from the home matrix the contacts of persons aged 70 and older to create an “elderly matrix”, because of the possible implementation of control measures on this specific population, especially in medicalized retirement homes. The rows and columns thus selected in the home contact matrix were set to 0, not to change the total amount of contacts across the five matrices.

#### 3.3.3 Estimation of hospital requirements and deaths

Based on the estimated number of new infected hospitalized cases per day provided by our transmission model, we inferred outcomes related to hospital requirements, namely hospitalization and ICU beds (Figure S2 and Table S1). First, we computed the number of cases which will require admission to ICU using age-dependent risks from the ‘Assistance Publique des Hôpitaux de Paris’ (AP-HP) in Paris, France (personal communication from P-Y. Boëlle). We then estimated the requirements in both non-ICU and ICU beds, considering the several age-specific lengths of stay observed in the AP-HP hospitals (without admission in ICU, pre-ICU and post-ICU admissions). Regarding the data at our disposal, we chose to only estimate the deaths of hospitalized infected cases, and we did not consider deaths in nursing homes and at home. To do this, we used the corresponding age-dependent risks of death and length of stay before death (set to be equal for both non-ICU and ICU) and estimated a weighting factor to match the observed number of deaths.

### 3.4 Estimation of transmission parameters

As mentionned before, the COVID-19 epidemic did not have a similar dynamic among the 13 metropolitan French Regions and regional healthcare facilities were not solicited at the same point. Therefore, to fit these differences and obtain better regional forecasts, we realized two successive parameter estimations using maximum likelihood estimation on the period stretching from March 20 (hereafter called *t*_1_) to March 28 (hereafter called *t*_9_), 2020. This period was chosen as data were available without missing value and still corresponded to the evolution of the epidemic before the implementation of the national lockdown.

First, we estimate the regional *β* parameter, governing the value of *R*_0_, and the initial lag, corresponding to the introduction date of SARS-CoV-2 in a specific region. We jointly estimated these two parameters by fitting both the daily number of hospitalizations approximated by a local polynomial regression (hereafter denoted *Hosp*) and the occupation of hospitalization beds (*BedHosp*) using the likelihood *L*_*β*_ defined in Equation (1).

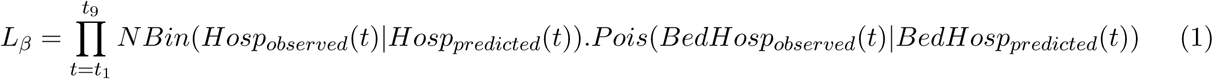

where *NBin*(.| *X*) is a negative binomial distribution of mean *X* and overdispersion 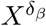, *δ*_*β*_ being a parameter specific to each region to be estimated, and *P ois*(.|*X*) is a Poisson distribution of lambda parameter *X*.

In a second step, we estimated three parameters: a multiplicative coefficient of the risk of ICU admission, a multiplicative coefficient of lengths of stay in ICU and the length of stay before death (equal between hospitalization and ICU) (Table S2). We jointly estimated these parameters by fitting the daily number of admissions in ICU and the number of deaths both approximated by a local polynomial regression (hereafter respectively denoted *ICU* and *Deaths*), and the occupation of ICU beds (*BedICU*) using the likelihood *L*_*ICU*_ defined in Equation (2).

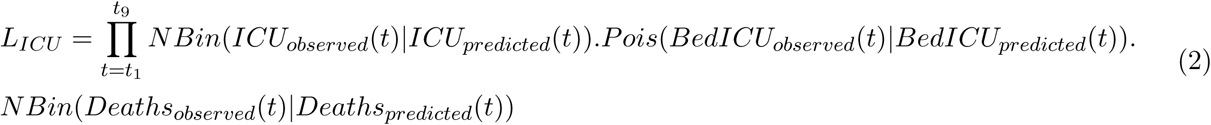

where *NBin*(.| *X*) is a negative binomial distribution of mean *X* and overdispersion 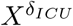 or 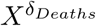 depending on the variable *X, δ*_*ICU*_ and *δ*_*Deaths*_ being parameters specific to each region to be estimated, and *P ois*(.|*X*) is a Poisson distribution of lambda parameter *X*.

The transmission model was implemented in C++. Data collection, data management, simulations, and results analysis and reporting were performed using R^17^ and the bbmle package.^18^

## 4 Results

### 4.1 Estimation of the pre-lockdown epidemiological dynamic

The first step of our analysis aims at reproducing the dynamic of the SARS-CoV-2 epidemic in France during the pre-lockdown stage. Figure 2 and Figures S3 to S14 illustrate the quality of the model predictions regarding hospital and ICU admissions, required beds, and deaths per day over the period of March 20 to 28, when the impact of the lockdown was still unseen on those outcomes. From our inference of the transmission probabilities for each French metropolitan region, we also estimate values of “pre-lockdown” reproduction number (*R*) for each of them. Hence, the median “regional” reproduction number was 2.80 with a minimum of 1.65 and a maximum of 4.19 (Table 3).

**Table 3:** Number of predicted symptomatic cases without lockdown between March 19 and April 19 2020, by region.

| Region | Symptomatic | AR symptomatic | R |
| --- | --- | --- | --- |
| Ile-de-France | 4051700 | 32.3% | 2.81 |
| Auvergne-Rhone-Alpes | 2092400 | 27.1% | 3.11 |
| Nouvelle-Aquitaine | 1694800 | 27.7% | 4.08 |
| Grand-Est | 1263700 | 22.8% | 2.19 |
| Pays de la Loire | 1172900 | 33.8% | 4.19 |
| Occitanie | 904700 | 16.2% | 2.94 |
| Normandie | 791100 | 22.5% | 3.54 |
| PACA | 737900 | 14.3% | 2.56 |
| Hauts-de-France | 681500 | 12.3% | 2.31 |
| Centre-Val de Loire | 666300 | 27.5% | 3.78 |
| Bourgogne-Franche-Comte | 561300 | 17.9% | 2.53 |
| Bretagne | 154100 | 4.6% | 2.22 |
| Corse | 25400 | 7.6% | 1.65 |
| **France** | **14797900** | **22.9%** | **-** |

AR: Attack rate

**Figure 2:**
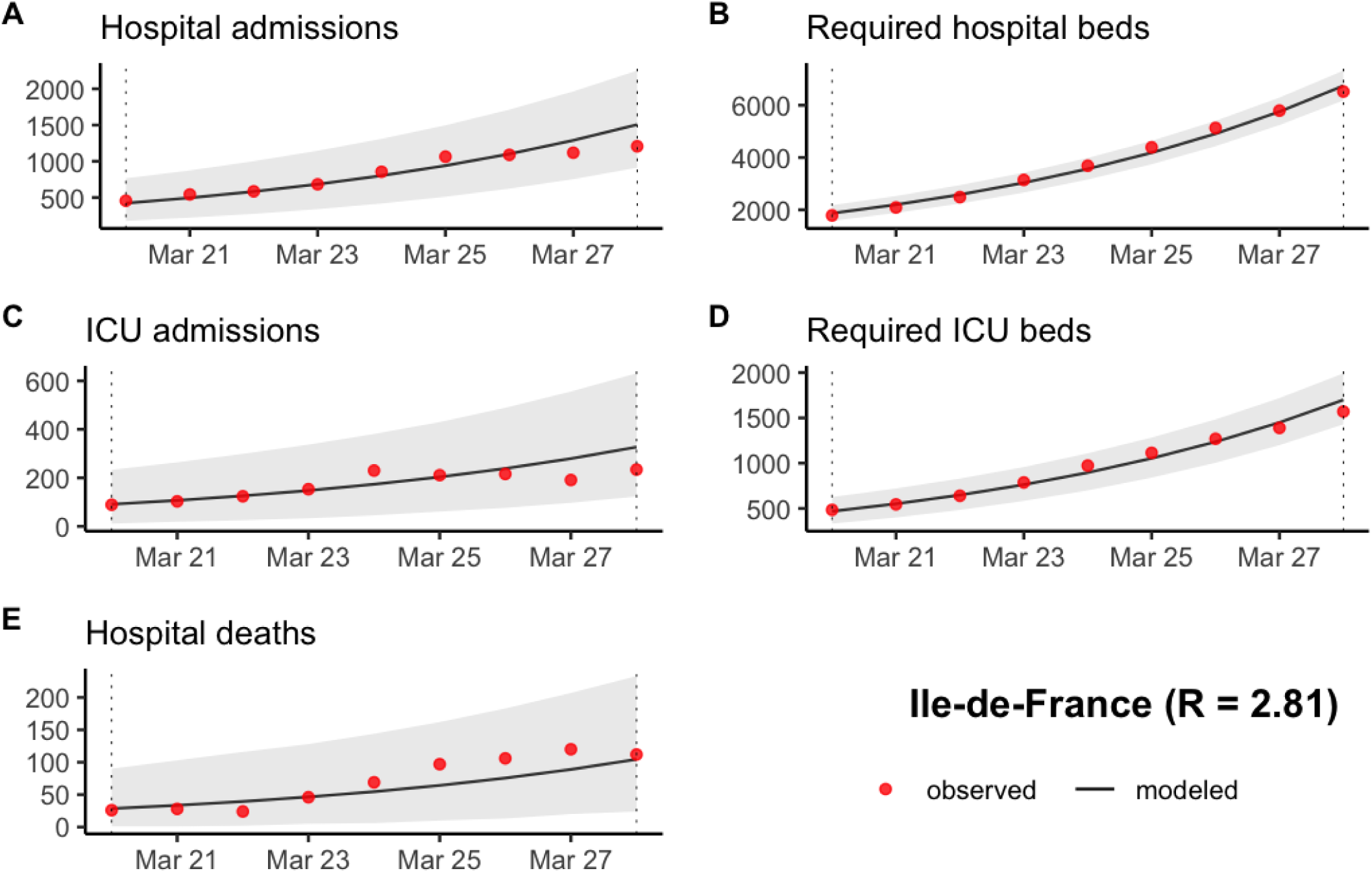
Prediction of (A) the number of new hospitalizations, (B) the number of required hospitalization beds, (C) the number of new ICU admissions, (D) the number of required ICU beds and (E) the number of new hospital deaths in Ile-de-France between March 20 and March 28 2020. The grey ribbon represents the 95% prediction interval.

### 4.2 Prevented epidemiological burden and hospital requirements averted

As illustrated by Figure S15 and Figure 3, our analysis shows that the number of new hospitalizations, and bed requirements, in every French Regions would have risen at an exponential rate with relatively minor geographic variations. However, all regions did not experience the same kind of introduction events (large clusters, superspreading events, etc.), hence the starting point of their epidemic was different and the lockdown occurred at different stages in each region depending on those initial factors.

**Figure 3:**
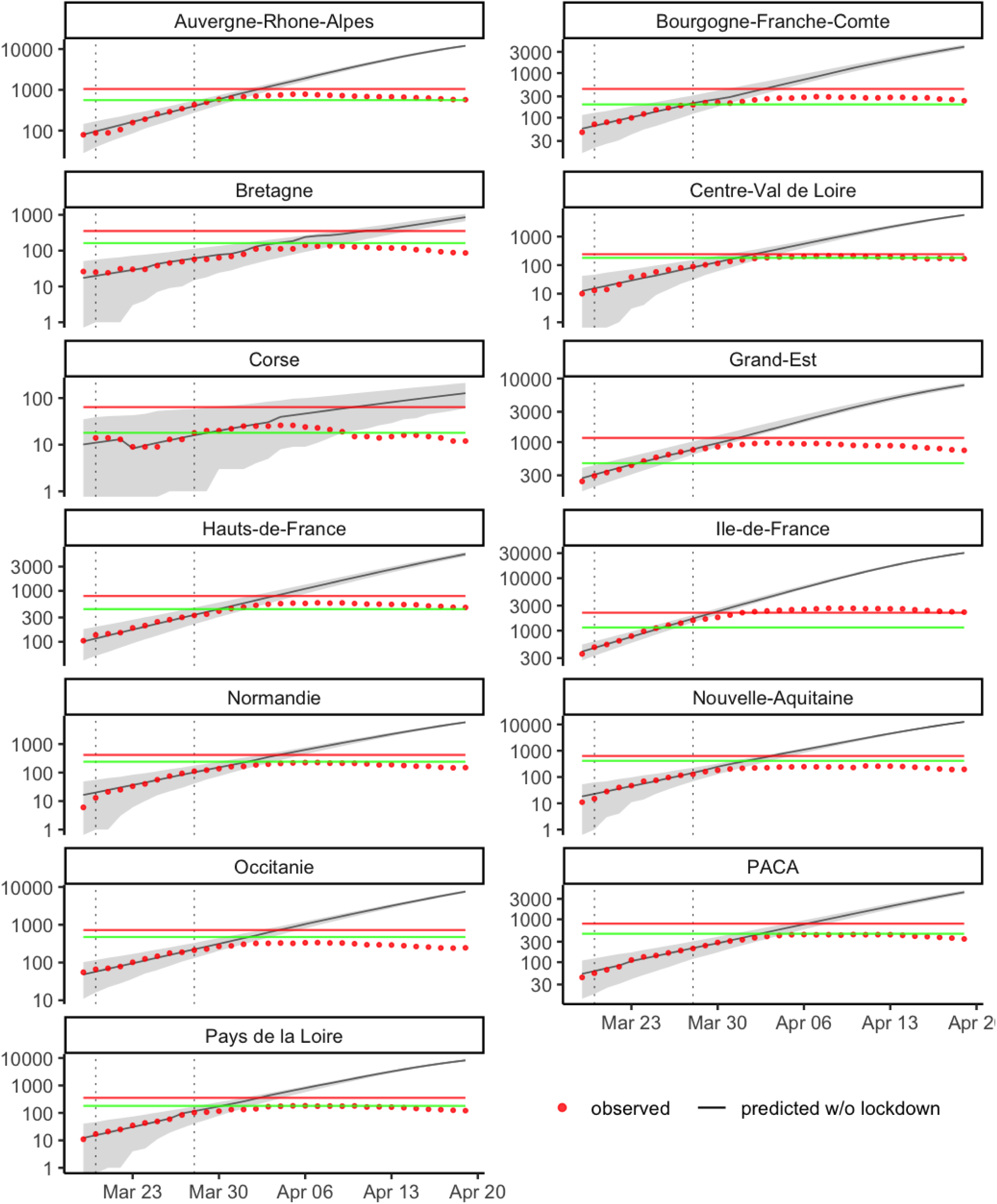
Estimation of the evolution of the number of ICU beds required by French metropolitan region without lockdown. The grey ribbon represents the 95% prediction interval. The red and green lines stand for the maximum number of ICU beds on April 2 2020 and the baseline maximum number of ICU beds, respectively.

These variations are also evidenced by the proportion of symptomatic infections by region. These proportions ranged from 32.3% in Ile-de-France to 4.6% in Bretagne. At the national level, we estimate that 22.9% of the French population would have experienced an episode of COVID-19 during the period of interest.

Hence, we notice in Table 4 that Ile-de-France benefited the most from the strategy, which averted 160,000 hospitalizations, 40,000 ICU admissions and the need for 27,600 ICU beds. At the other end of the spectrum, the lockdown in Corse prevented less than 900 hospitalizations and the need for 100 additional ICU beds, which still represent a reduction of 71.5% and 76.9% of the expected burden. Although most

**Table 4:** Epidemiological outcomes and hospital requirements observed with current lockdown, and predicted without lockdown between March 19 and April 19 2020, by region.

| Region | Hospitalizations |  |  |  | ICU admissions |  |  |  | Required ICU beds* |  |  |  |
| --- | --- | --- | --- | --- | --- | --- | --- | --- | --- | --- | --- | --- |
|  | No lockdown | Observed | Difference | Rel. Reduction | No lockdown | Observed | Difference | Rel. Reduction | No lockdown | Observed | Difference | Rel. Reduction |
| Ile-de-France | 193620 [149200-244650] | 31040 | -162580 | -84% | 45420 [28420-67000] | 5420 | -40010 | -88.1% | 30280 [29100-31480] | 2670 | -27610 | -91.2% |
| Auvergne-Rhone-Alpes | 94640 [77120-114560] | 8190 | -86450 | -91.3% | 21100 [15210-28390] | 1530 | -19560 | -92.7% | 12030 [11290-12780] | 780 | -11240 | -93.4% |
| Nouvelle-Aquitaine | 72920 [40000-116740] | 2560 | -70360 | -96.5% | 16710 [10300-25260] | 500 | -16200 | -96.9% | 12540 [11780-13320] | 260 | -12280 | -97.9% |
| Pays de la Loire | 52670 [41820-65420] | 2060 | -50620 | -96.1% | 11170 [7410-16200] | 350 | -10820 | -96.9% | 8260 [7640-8890] | 180 | -8080 | -97.8% |
| Grand-Est | 62980 [38840-93460] | 13230 | -49750 | -79% | 14420 [6000-26780] | 2030 | -12390 | -85.9% | 7900 [7300-8510] | 970 | -6930 | -87.7% |
| Occitanie | 37260 [27760-48710] | 3010 | -34250 | -91.9% | 9130 [5550-14060] | 700 | -8420 | -92.2% | 7570 [6980-8180] | 340 | -7240 | -95.6% |
| Normandie | 32710 [24370-42870] | 1830 | -30880 | -94.4% | 7870 [4760-12260] | 400 | -7470 | -94.9% | 5800 [5290-6330] | 220 | -5580 | -96.2% |
| Centre-Val de Loire | 29000 [21300-38480] | 2230 | -26770 | -92.3% | 7010 [4130-11110] | 400 | -6600 | -94.2% | 5740 [5230-6260] | 210 | -5520 | -96.2% |
| PACA | 31590 [21330-44380] | 5000 | -26590 | -84.2% | 6290 [3170-10900] | 850 | -5440 | -86.5% | 4300 [3860-4750] | 440 | -3850 | -89.5% |
| Hauts-de-France | 29280 [19470-41510] | 6620 | -22660 | -77.4% | 7480 [4020-12390] | 1110 | -6370 | -85.2% | 5260 [4770-5770] | 580 | -4680 | -89% |
| Bourgogne-Franche-Comte | 25220 [10210-47400] | 3930 | -21290 | -84.4% | 6130 [2060-12730] | 570 | -5560 | -90.7% | 3890 [3470-4330] | 300 | -3600 | -92.5% |
| Bretagne | 6330 [2970-11260] | 1240 | -5080 | -80.3% | 1600 [340-4100] | 250 | -1360 | -85% | 850 [660-1060] | 140 | -710 | -83.5% |
| Corse | 1230 [40-4090] | 360 | -880 | -71.5% | 240 [0-1310] | 50 | -190 | -79.2% | 130 [60-210] | 30 | -100 | -76.9% |
| **Total** | **669470 [474430-913520]** | **81740** | **-587730** | **-87.8%** | **154570 [91360-242500]** | **14250** | **-140320** | **-90.8%** | **104550 [97430-111880]** | **7180** | **-97380** | **-93.1%** |

\* Maximum number of ICU beds occupied at one point over the period

French regions experienced tensions on their healthcare system, our analysis shows that the weight of COVID-19 cases, with the lockdown, was only a fraction (from 4% to 30%) of what they would have had to manage without the strategy.

Over all Metropolitan France, we predict that almost 670,000 patients would have been hospitalized between March 19 and April 19, and more than 100,000 ICU beds would have been required for the care of 155,000 patients needing ICU attention.

Regarding the mortality burden in hospitals, its evolution without lockdown is predicted to follow the same path of the previously estimated outcomes (Figure S16). Hence, we estimate that almost 74,000 patients would have died in hospitals over the same period, and that the lockdown strategy reduced this burden by 83.5%, to 12,000 deaths (Table 5).

**Table 5:**
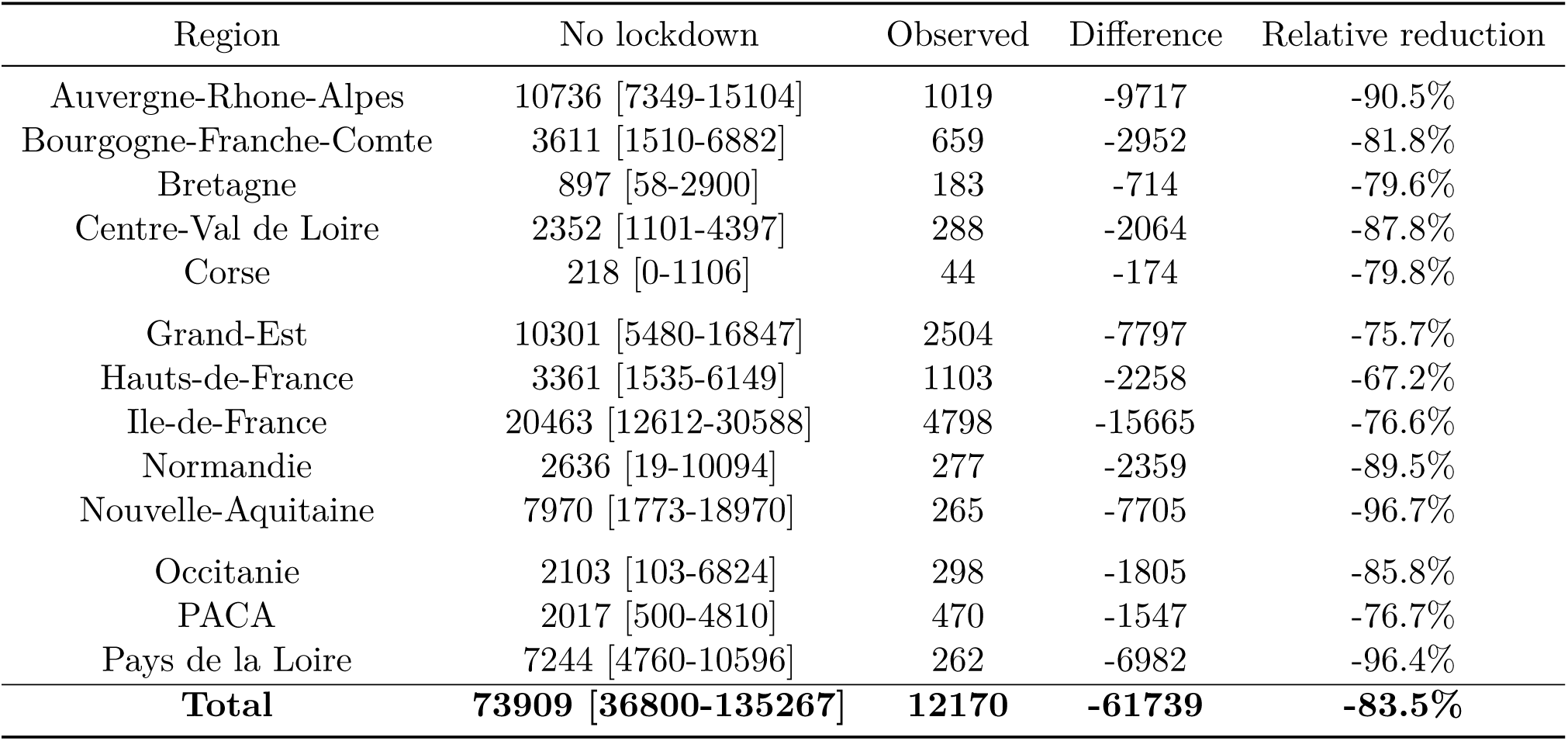
Number of hospital deaths observed with current lockdown, and predicted without lockdown between March 19 and April 19 2020, by region.

| Region | No lockdown | Observed | Difference | Relative reduction |
| --- | --- | --- | --- | --- |
| Auvergne-Rhone-Alpes | 10736 [7349-15104] | 1019 | -9717 | -90.5% |
| Bourgogne-Franche-Comte | 3611 [1510-6882] | 659 | -2952 | -81.8% |
| Bretagne | 897 [58-2900] | 183 | -714 | -79.6% |
| Centre-Val de Loire | 2352 [1101-4397] | 288 | -2064 | -87.8% |
| Corse | 218 [0-1106] | 44 | -174 | -79.8% |
| Grand-Est | 10301 [5480-16847] | 2504 | -7797 | -75.7% |
| Hauts-de-France | 3361 [1535-6149] | 1103 | -2258 | -67.2% |
| Ile-de-France | 20463 [12612-30588] | 4798 | -15665 | -76.6% |
| Normandie | 2636 [19-10094] | 277 | -2359 | -89.5% |
| Nouvelle-Aquitaine | 7970 [1773-18970] | 265 | -7705 | -96.7% |
| Occitanie | 2103 [103-6824] | 298 | -1805 | -85.8% |
| PACA | 2017 [500-4810] | 470 | -1547 | -76.7% |
| Pays de la Loire | 7244 [4760-10596] | 262 | -6982 | -96.4% |
| **Total** | **73909 [36800-135267]** | **12170** | **-61739** | **-83.5%** |

## 5 Discussion

The purpose of this study was to assess the regional and national impacts of the French lockdown on epidemiological outcomes, namely hospital requirements and mortality, using a deterministic, compartmental model. Indeed, one month after its beginning, to our knowledge, no figures are available on the averted burden of the COVID-19 in France.

From March 19 to April 19, we estimated that the lockdown might have averted around 590,000 hospitalizations and 140,000 ICU admissions over all metropolitan France, corresponding to relative reductions of 88% and 91%, respectively. We estimated that, over this period, more than 100,000 ICU beds might have been occupied at the same time at the national level. Regarding the mortality burden of the COVID-19, the French lockdown might have averted slightly more than 61,000 deaths in hospitals. These results varied slightly between regions, but the lockdown seems to have had an important impact in all regions. Indeed, the regional relative reductions ranged from 70% to 98% for the different considered hospital requirements and deaths. Related to theses regional disparities, we noted that the estimated reproduction numbers were surprisingly high in some regions, especially in Pays de la Loire and Nouvelle-Aquitaine with values greater than 4. However, these values may not reflect a high number of cases in these regions, but may be likely related to a more recent introduction of COVID-19 in the region (smaller initial lag were estimated for these regions compared to the other regions). This may also be due to population movements before the start of the lockdown to these two specific regions located on the West coast of France, which could contribute to bias our estimates. However, this assumption should be confirmed with geolocalisation of mobile phones for example, as data currently at our disposal do not allow us to answer this question.

Taking into account the latest information on the SARS-CoV-2 and its characteristics, namely regarding its transmission and its infectivity, the model used here is an enhanced version of our previous model developed to forecast hospital needs at the start of the epidemic in France.^5^ The present model is composed of 8 compartments, whom 5 are dedicated to the infected phase, compared to 1 in a classic SEIR model. In more details, based on the knowledge on the transmission of the virus, we included two compartements (pre-symptomatic and asymptomatic) dedicated to cases without symptoms and being contagious. Moreover, we used up-to-date transmission parameters’ values, such as the 90% decrease of infectivity for asymptomatics compared to symptomatics cases.^15^ Regarding the computation of outcomes of interest (hospital requirements and deaths), we considered the whole care pathway of patients inside hospitals, taking into account the pre- and post-ICU hospitalizations for cases going into ICU. This holistic consideration leads to a precise quantification of the required beds to be as close as possible to the observed bed occupancy. Moreover, these estimations were based on observed lengths of stay and risks of ICU admissions in real settings and modulated for each region with specific estimated coefficients to account for regional disparities. In addition, we estimated the differences and relative reductions of outcomes based on the national daily data on the epidemic evolution, gathered by SpF, which allows to provide an accurate quantification of the averted burden.

We estimated our transmission parameters from data on hospitalizations, ICU admissions, and deaths between March 20 and March 28. Therefore, the differences and reductions presented here are relative to what would have happened if the transmission dynamics had remained unchanged. We did not model what impact could have had moderate mitigation measures compared to the lockdown. However, several studies already estimated that these moderate measures would be largely insufficient at mitigating the epidemic.^8,9^ The impact of seasonal variations in the transmission of SARS-Cov-2 is still unclear; therefore, we chose to not include a seasonal parameter in our model. Although data on the matter is very scarce, several experts suggest that the impact of seasonality on COVID-19 transmission could be modest.^19,20^ It appears thus reasonable to assume that the vast majority of the reduction has to be credited to the lockdown, even if it is impossible, for the moment, to disentangle the effect of the lockdown and the potential underlying effect of seasonality. Moreover, the mortality estimates presented here do not take into account deaths occurring outside hospitals (at home or in nursing homes) as we limited our analysis to hospital data. As with many countries, the true overall mortality in France remains unknown, as it is likely that many deaths actually related to COVID-19 might not have been identified as such. The impact of the lockdown may thus remain unknown regarding overall mortality. Our model also does not account for the excess mortality that would have resulted from hospital congestion. Given the important excess hospitalizations and ICU admissions that would have occurred without the lockdown, it is likely that it would have resulted in an important excess mortality due to the congestion of the health care system. Another point of concern is the underestimation of the number of deaths that would have occurred without the lockdown as the analysis was cutoff at one month, while a proportion of patients still hospitalized on April 19 might die afterwards, given the delay between hospital admission and death. Therefore, the lockdown might have averted more deaths overall than what is estimated here. Furthermore, our model does not include population movements between regions. At the time, we assumed that the impact would be negligible, as infected cases were present in most locations, meaning that the transmission was mainly driven locally. It is even more the case during the lockdown as travels are greatly reduced. Finally, we did not take into account the fact that some regions (like Grand-Est or Hauts-de-France) implemented mitigation strategies before the national lockdown because of clusters of infected persons. As previously discussed, the impact of such measures was likely to be modest at the time, and even so, this bias would tend to minimize the estimated impact of the lockdown. However, we can consider these measures were taken into account in the estimated reproduction number.

The results presented here, suggesting an important impact of the lockdown, are consistent with other studies. Flaxman and colleagues, used a semi-mechanistic Bayesian hierarchical model to attempt to infer the impact of control interventions across 11 European countries, including France.^21^ They estimated that the lockdown managed to reduce the reproduction number below one, and that, up to March 31 only, up to 4,800 deaths might have been averted (56% relative reduction). Salje and colleagues used an age-stratified, deterministic, compartmental model to assess the impact of the lockdown in France, and current population immunity.^22^ They estimated that the first month of the lockdown reduced the reproduction number from 3.3 to 0.5 (84% reduction). This important reduction of is consistent with our results, suggesting a high effectiveness of the lockdown in France. They also estimated that on May 11, the currently scheduled date for exiting the lockdown, between 3.5% and 10.3% of the population will have been infected. Their estimations are almost identical for April 19 (around 3.4% to 10.1%). On the same day, we estimated that, without the lockdown, the attack rate of symptomatic cases would have been around 23%. Although we should also take into account asymptomatic cases, it is likely that without the lockdown, herd immunity might still not have been achieved at this point. Hence, the one-month burden we quantify underestimates the total burden of an un-controlled COVID-19 epidemic. Consequently, our results highlight the possible life cost of letting the COVID-19 epidemic run its course, and the need for various combinations of control measures to avoid a second wave of the epidemic after exiting the lockdown. Di Domenico and colleagues investigated possible exit strategies using a stochastic, age-structured, transmission model integrating data on age profile and social contacts in the Île-de-France region.^23^ They also estimated that exiting the lockdown completely without other control measures would lead to a second wave largely overwhelming the healthcare system.

However, there is still a significant number of unknowns regarding the physiopathology of COVID-19. For instance, how age influences the susceptibility to infection, the probability of developing an asymptomatic form, the infectiousness of asymptomatic cases. Moreover, the impact of other factors on the transmission of SARS-CoV-2 (climatic factors, seasonal variations) remains unclear. Consequently, the future course of the epidemic is still subject to many uncertainties. More work is still needed to forecast the behavior of the epidemic under various scenarios for the weeks to come.

## 6 Conclusion

With a month’s hindsight, our study shows that the national lockdown, started in France on March 17, 2020, had an important impact on the evolution of the COVID-19 epidemic. This unprecedented measure drastically reduced the number of hospital and ICU admissions, and prevented a large number of deaths at the national level. Without this lockdown, the COVID-19 epidemic would have had a critical morbidity and mortality burden in France, overwhelming the French healthcare system in a matter of weeks. Without any efficient treatment or vaccine, our results reinforce the need to control the COVID-19 pandemic by measures at least as efficient as a nation-wide lockdown.

## Data Availability

Source code is available upon request.

## 7 Acknowlegments

The authors would like to thank Simon Cauchemez and Pierre-Yves Boëlle for their fruitful insights. This work is partly funded by the ANR Project SPHINx.

## 9 Supplementary material

**Figure S1:**
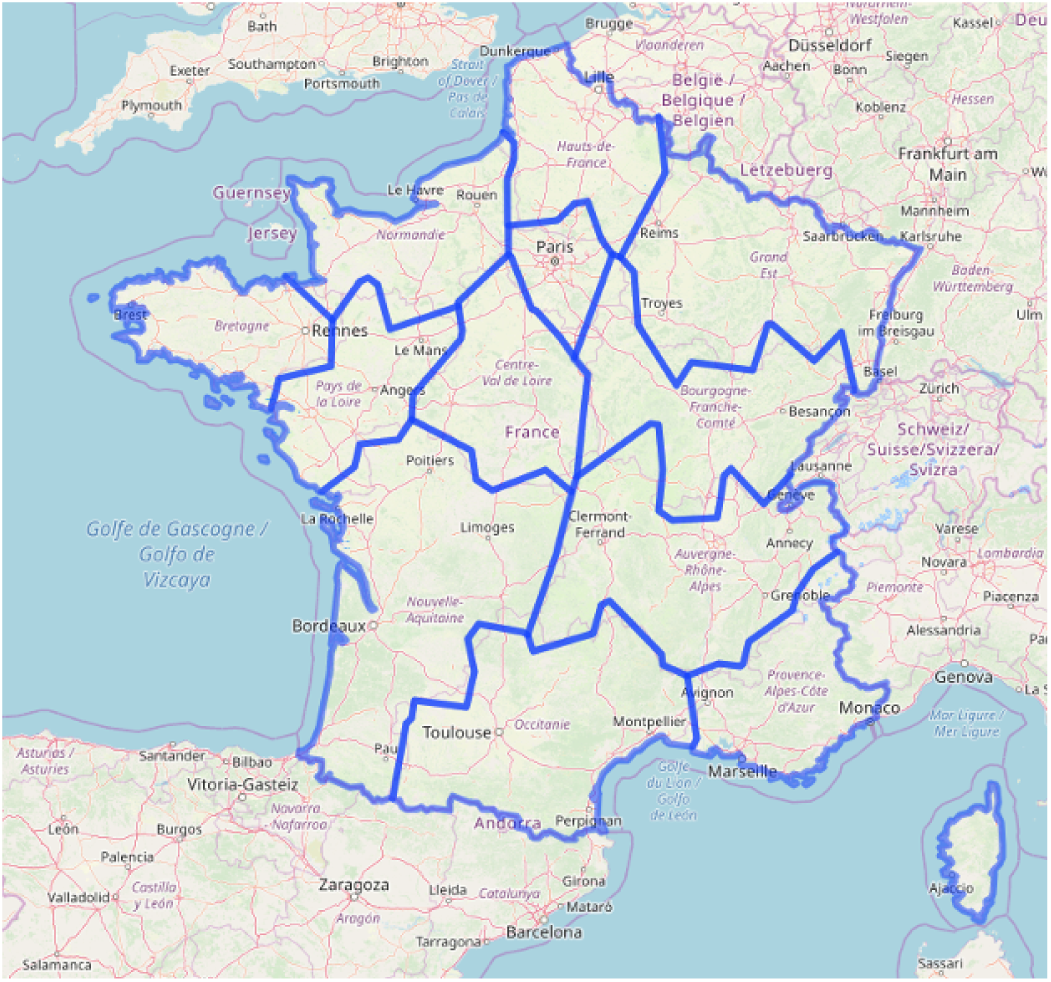
Map of France divided into its 13 Regional catchment areas.

**Table S1:** Mean lengths of stay used to compute the number of required beds by region.

| Region | Hospitalization los | ICU los* | Pre-ICU los | Post-ICU los |
| --- | --- | --- | --- | --- |
| Auvergne-Rhone-Alpes | 13.66 | 6.13 | 1.38 | 7.80 |
| Hauts-de-France | 13.29 | 12.16 | 1.37 | 7.87 |
| PACA | 13.24 | 7.81 | 1.38 | 7.82 |
| Grand-Est | 13.49 | 10.08 | 1.37 | 7.82 |
| Occitanie | 13.51 | 12.77 | 1.38 | 7.86 |
| Normandie | 13.67 | 8.48 | 1.37 | 7.80 |
| Nouvelle-Aquitaine | 13.94 | 10.06 | 1.38 | 7.88 |
| Centre-Val de Loire | 13.91 | 12.69 | 1.38 | 7.86 |
| Bourgogne-Franche-Comte | 13.51 | 10.45 | 1.37 | 7.87 |
| Bretagne | 13.16 | 6.84 | 1.37 | 7.82 |
| Pays de la Loire | 13.89 | 12.64 | 1.37 | 7.86 |
| Ile-de-France | 13.47 | 10.06 | 1.36 | 7.83 |
| Corse | 12.98 | 12.79 | 1.38 | 7.83 |

los = Length of stay
\*Coefficient associated to length of stay in ICU taken into account

**Figure S2:**
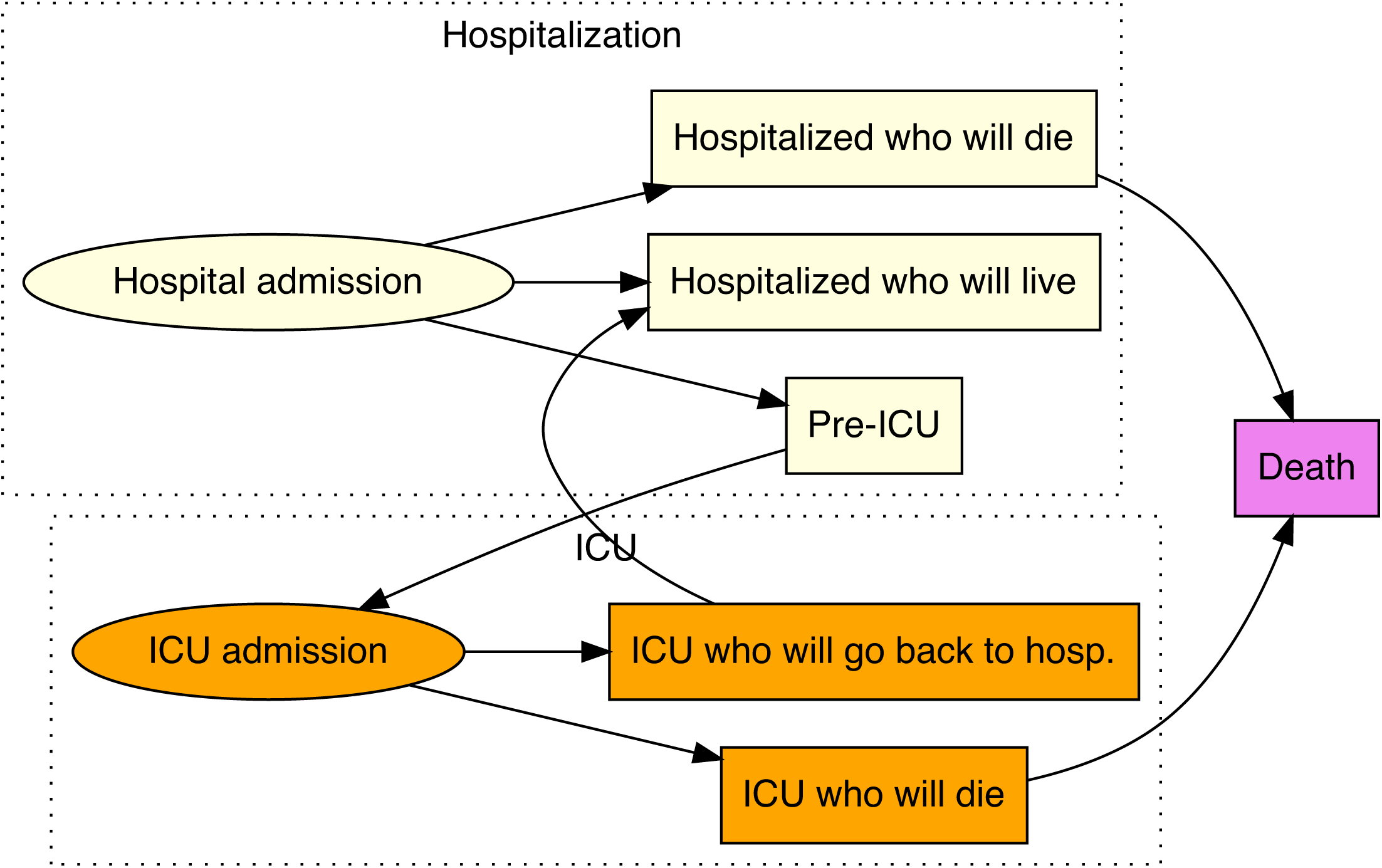
Diagram of hospitalized COVID-19 patient care pathways.

**Table S2:** Regional coefficients linked to risk of ICU admission, length of stay in ICU and length of stay between admission in hospitalization or ICU and deaths.

| Region | Coefficient associated to risk of ICU admission |  | Coefficient associated to length of stay in ICU |  | delta ICU | Length of stay between admission and death* |  |  |
| --- | --- | --- | --- | --- | --- | --- | --- | --- |
|  | Estimate | 95% Confidence Interval | Estimate | 95% Confidence Interval |  | Estimate | 95% Confidence Interval | delta Deaths |
| Auvergne-Rhone-Alpes | 0.89 | - | 0.49 | - | 1.00 | 6.50 | - | 1.00 |
| Hauts-de-France | 1.00 | [1.00-1.00] | 0.96 | [0.00-1.00] | 0.97 | 7.05 | [3.62-13.72] | 1.00 |
| PACA | 0.79 | [0.73-0.84] | 0.59 | [0.58-0.60] | 0.92 | 9.50 | [8.99-10.04] | 0.62 |
| Grand-Est | 0.87 | [0.83-0.90] | 0.81 | [0.79-0.83] | 0.55 | 5.50 | [5.39- 5.61] | 0.73 |
| Occitanie | 1.00 | [0.75-1.00] | 1.00 | - | 1.00 | 9.56 | [8.52-10.73] | 0.17 |
| Normandie | 1.00 | [0.04-1.00] | 0.67 | [0.65-0.69] | 1.00 | 6.66 | - | 0.00 |
| Nouvelle-Aquitaine | 0.96 | [0.81-0.99] | 0.79 | [0.77-0.81] | 0.73 | 5.49 | [5.11- 5.90] | 0.29 |
| Centre-Val de Loire | 1.00 | [1.00-1.00] | 0.99 | [0.99-0.99] | 1.00 | 7.49 | [7.20- 7.80] | 1.00 |
| Bourgogne-Franche-Comte | 0.96 | [0.84-0.99] | 0.81 | [0.79-0.82] | 0.58 | 5.23 | [2.22-12.30] | 0.81 |
| Bretagne | 1.00 | [0.00-1.00] | 0.56 | [0.56-0.57] | 0.88 | 5.50 | [5.14- 5.88] | 0.45 |
| Pays de la Loire | 0.87 | [0.77-0.93] | 1.00 | - | 1.00 | 4.21 | [1.21-14.62] | 1.00 |
| Ile-de-France | 0.90 | [0.87-0.92] | 0.79 | [0.78-0.80] | 0.62 | 7.97 | [5.00-12.70] | 0.67 |
| Corse | 0.72 | [0.61-0.80] | 1.00 | - | 1.00 | 3.63 | [1.07-12.30] | 1.00 |

Some 95% Confidence Interval could have not been computed and are represented by a dash
\*Either hospitalization or ICU

**Figure S3:**
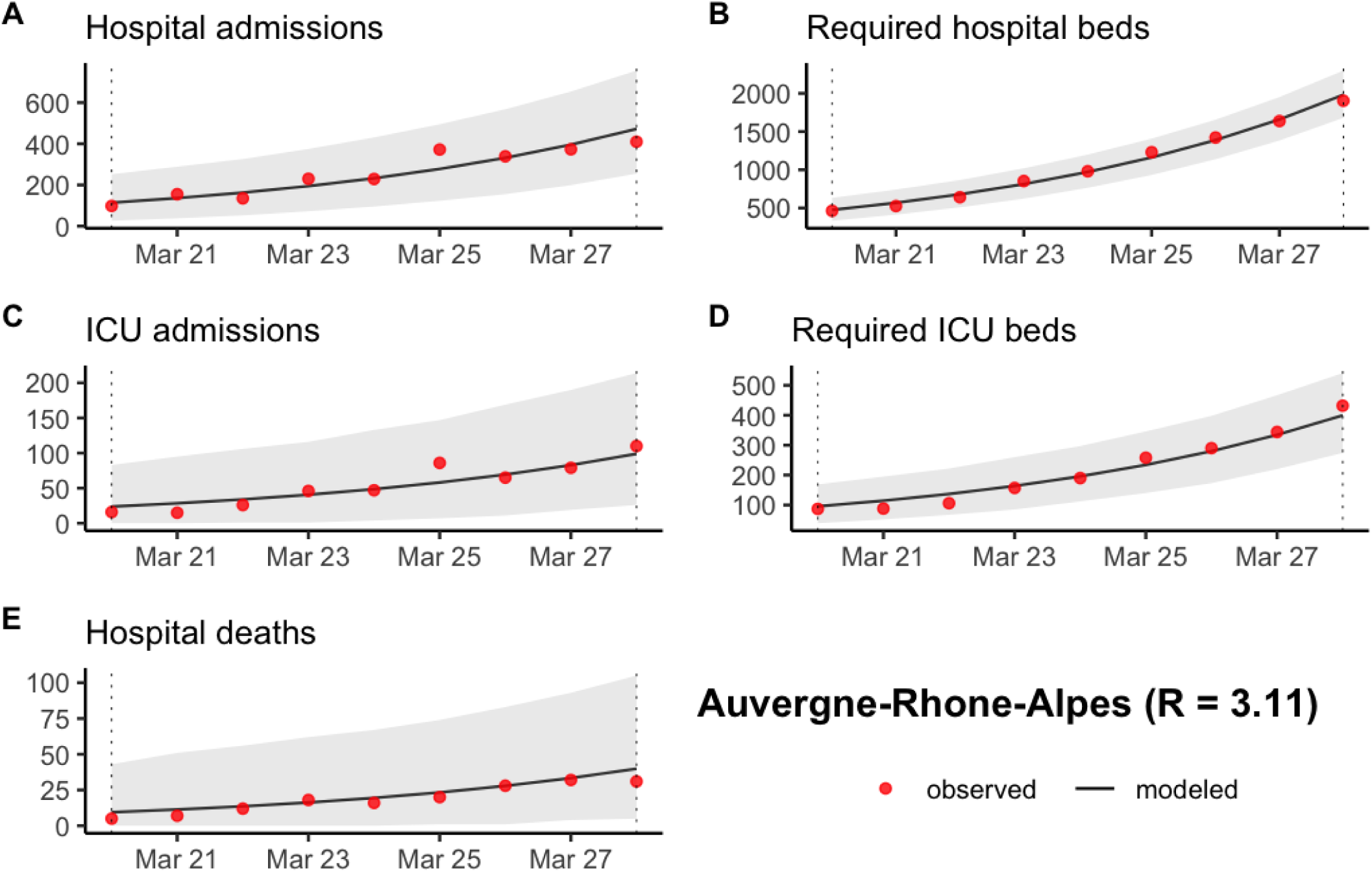
Prediction of (A) the number of new hospitalizations, (B) the number of required hospitalization beds, (C) the number of new ICU admissions, (D) the number of required ICU beds and (E) the number of new hospital deaths in Auvergne-Rhone-Alpes between March 20 and March 28 2020. The grey ribbon represents the 95% prediction interval.

**Figure S4:**
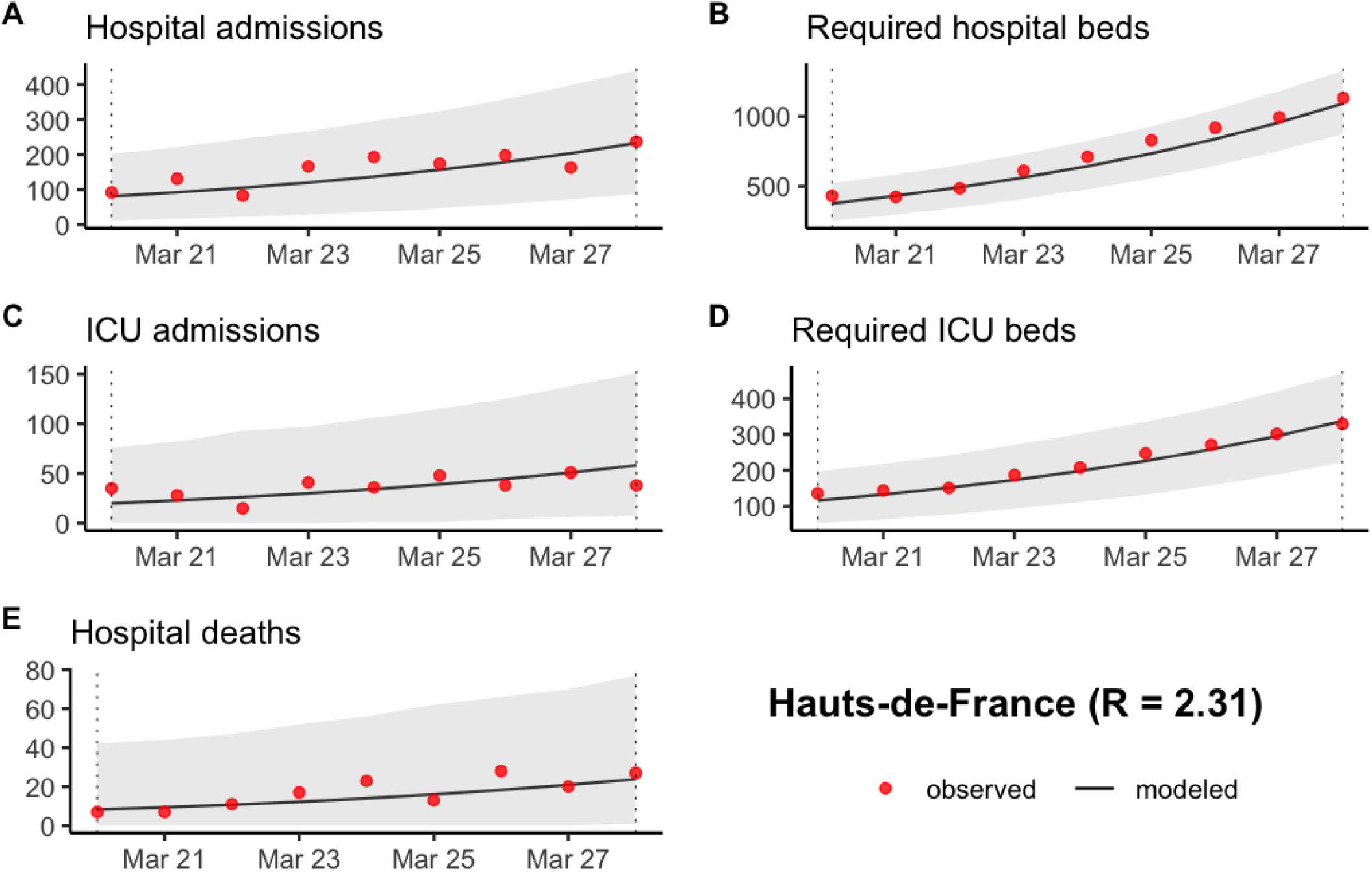
Prediction of (A) the number of new hospitalizations, (B) the number of required hospitalization beds, (C) the number of new ICU admissions, (D) the number of required ICU beds and (E) the number of new hospital deaths in Hauts-de-France between March 20 and March 28 2020. The grey ribbon represents the 95% prediction interval.

**Figure S5:**
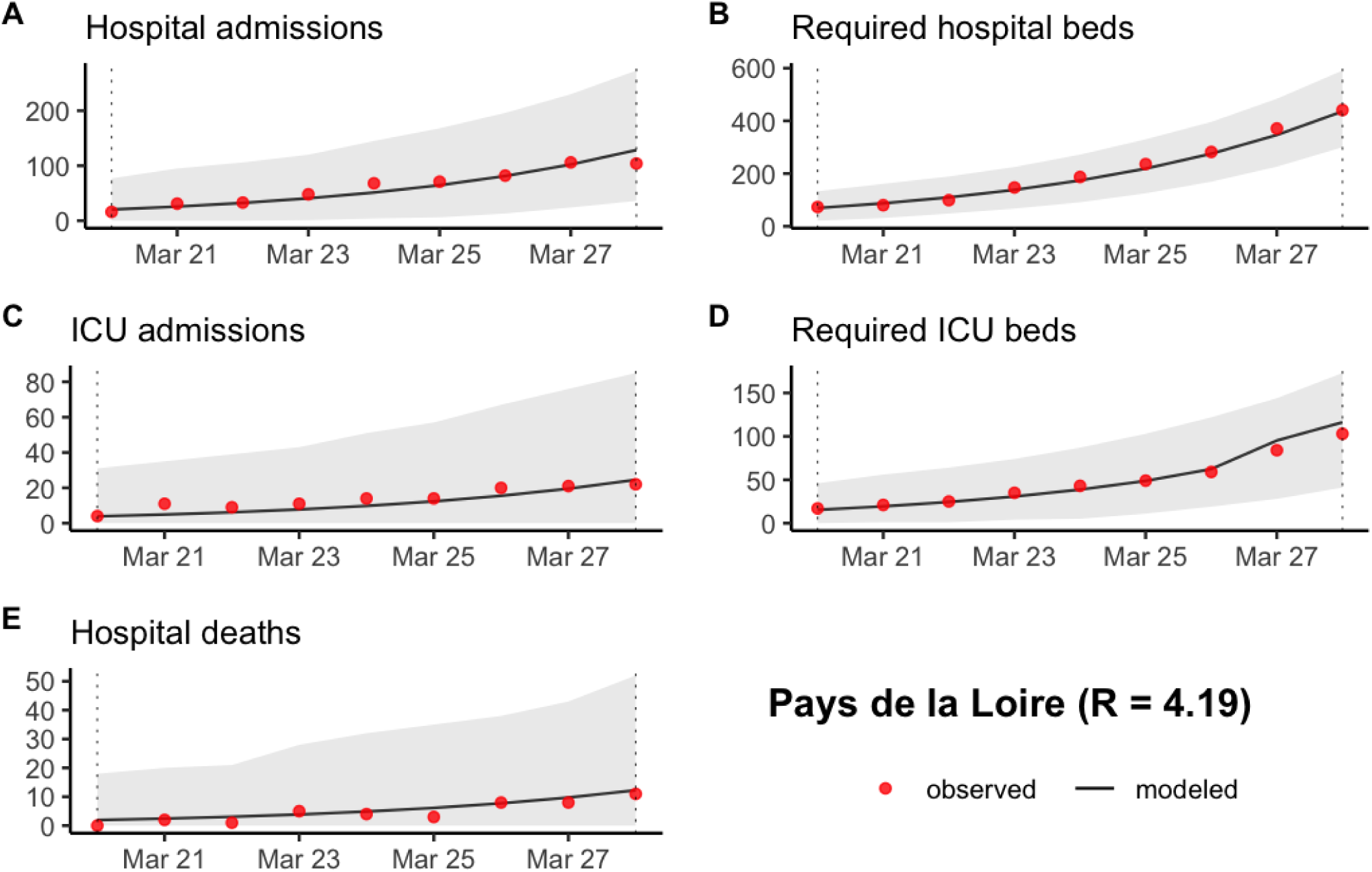
Prediction of (A) the number of new hospitalizations, (B) the number of required hospitalization beds, (C) the number of new ICU admissions, (D) the number of required ICU beds and (E) the number of new hospital deaths in Pays-de-la-Loire between March 20 and March 28 2020. The grey ribbon represents the 95% prediction interval.

**Figure S6:**
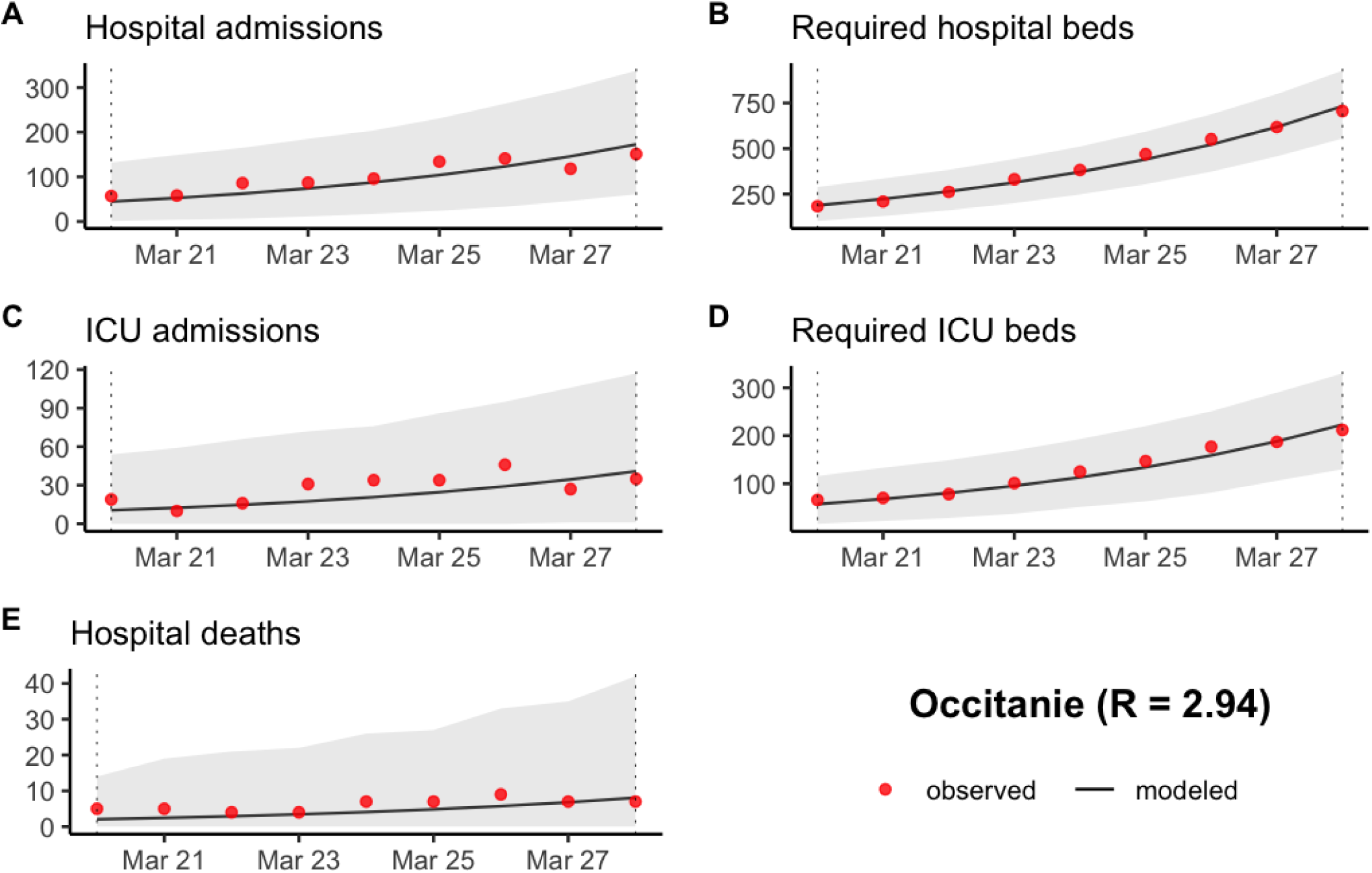
Prediction of (A) the number of new hospitalizations, (B) the number of required hospitalization beds, (C) the number of new ICU admissions, (D) the number of required ICU beds and (E) the number of new hospital deaths in Occitanie between March 20 and March 28 2020. The grey ribbon represents the 95% prediction interval.

**Figure S7:**
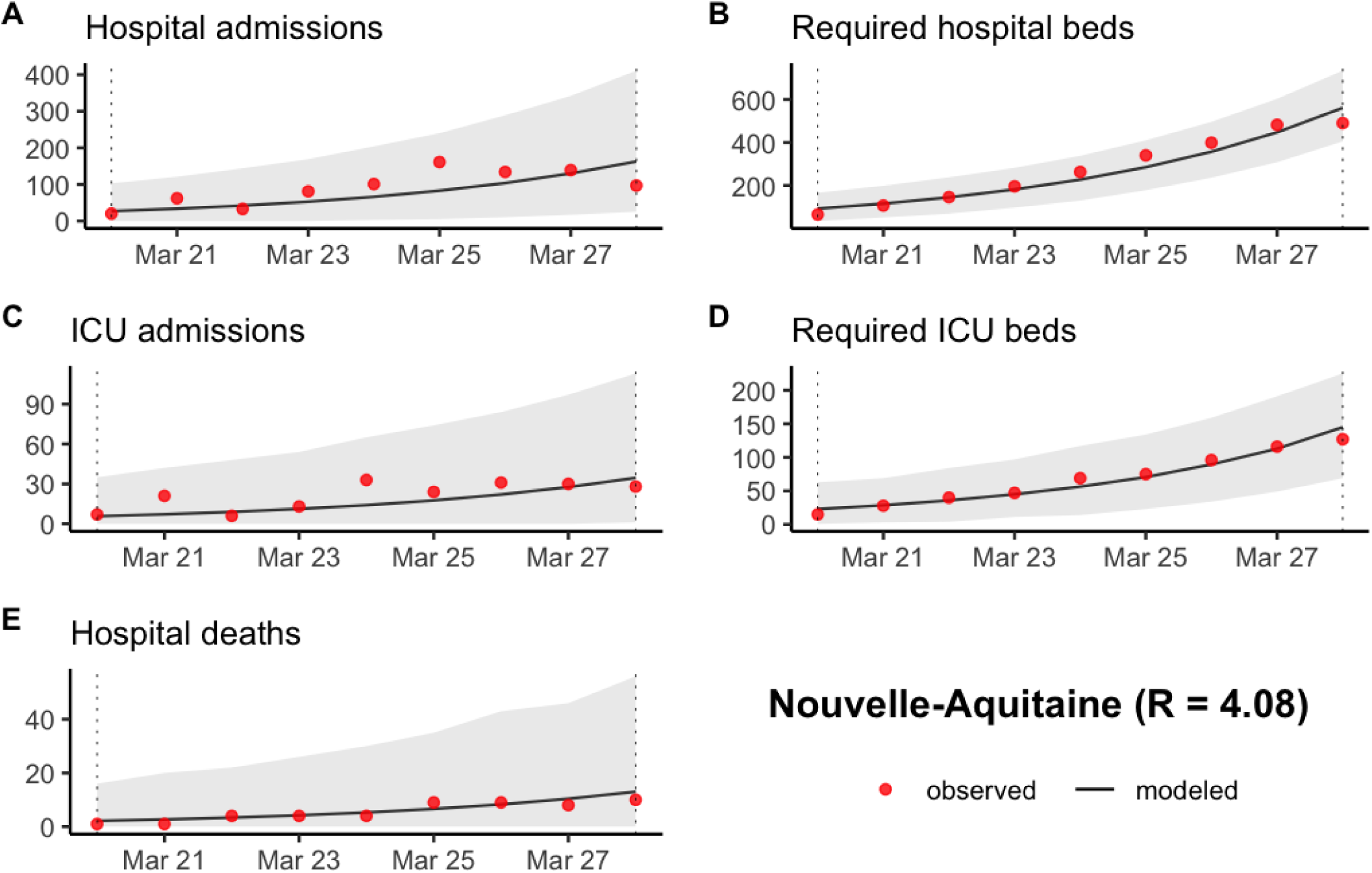
Prediction of (A) the number of new hospitalizations, (B) the number of required hospitalization beds, (C) the number of new ICU admissions, (D) the number of required ICU beds and (E) the number of new hospital deaths in Nouvelle-Aquitaine between March 20 and March 28 2020. The grey ribbon represents the 95% prediction interval.

**Figure S8:**
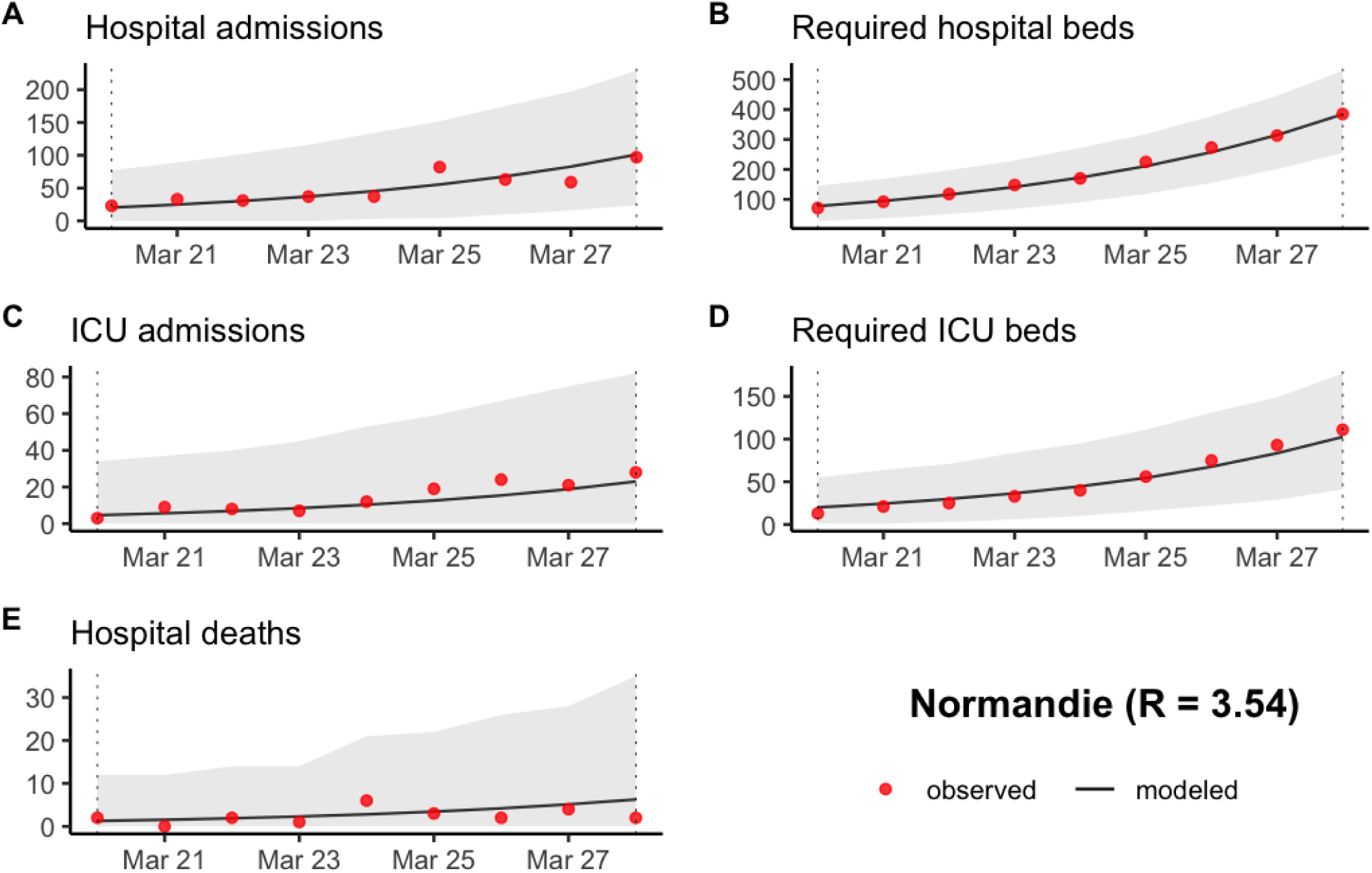
Prediction of (A) the number of new hospitalizations, (B) the number of required hospitalization beds, (C) the number of new ICU admissions, (D) the number of required ICU beds and (E) the number of new hospital deaths in Normandie between March 20 and March 28 2020. The grey ribbon represents the 95% prediction interval.

**Figure S9:**
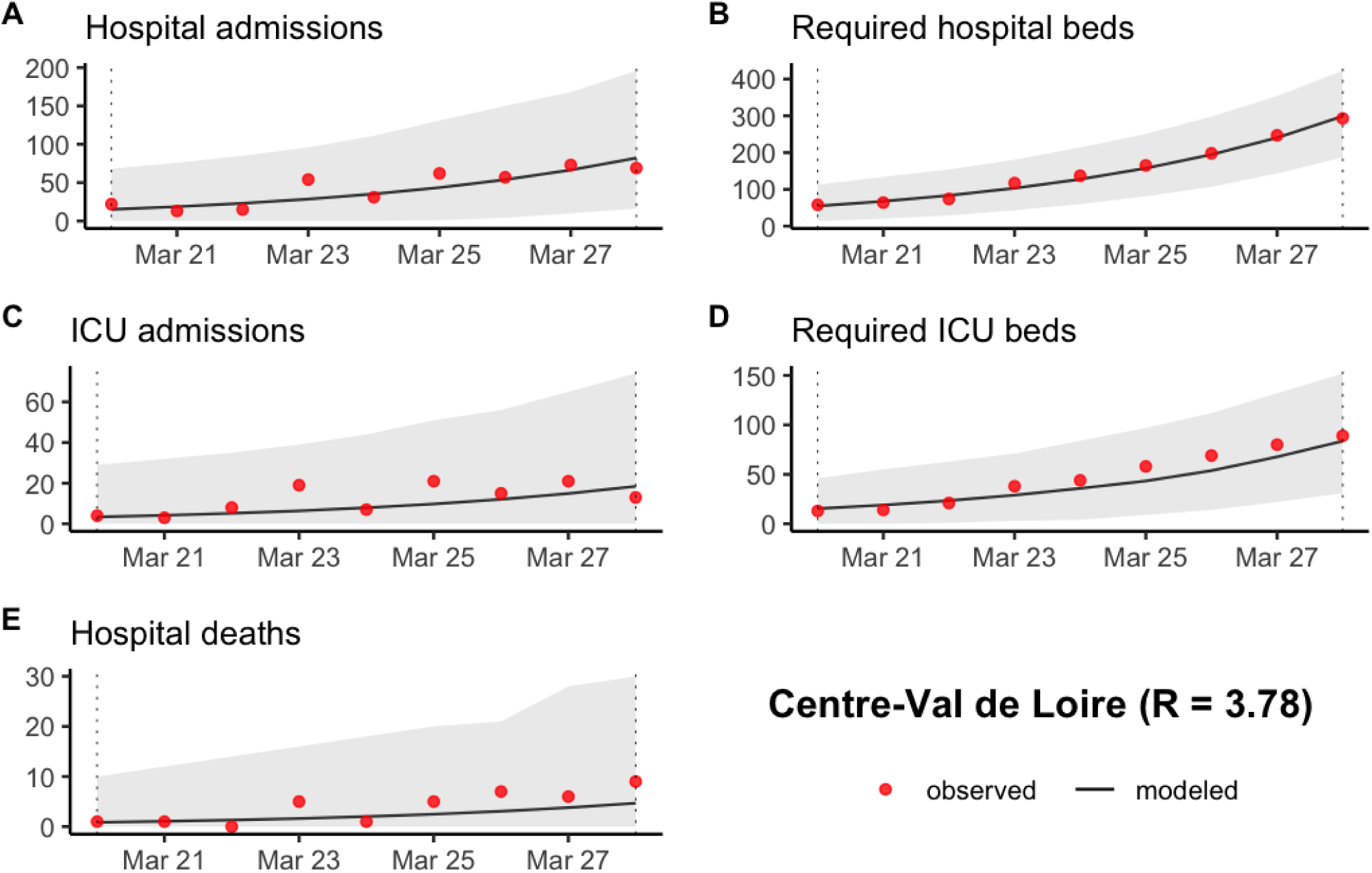
Prediction of (A) the number of new hospitalizations, (B) the number of required hospitalization beds, (C) the number of new ICU admissions, (D) the number of required ICU beds and (E) the number of new hospital deaths in Centre-Val de Loire between March 20 and March 28 2020. The grey ribbon represents the 95% prediction interval.

**Figure S10:**
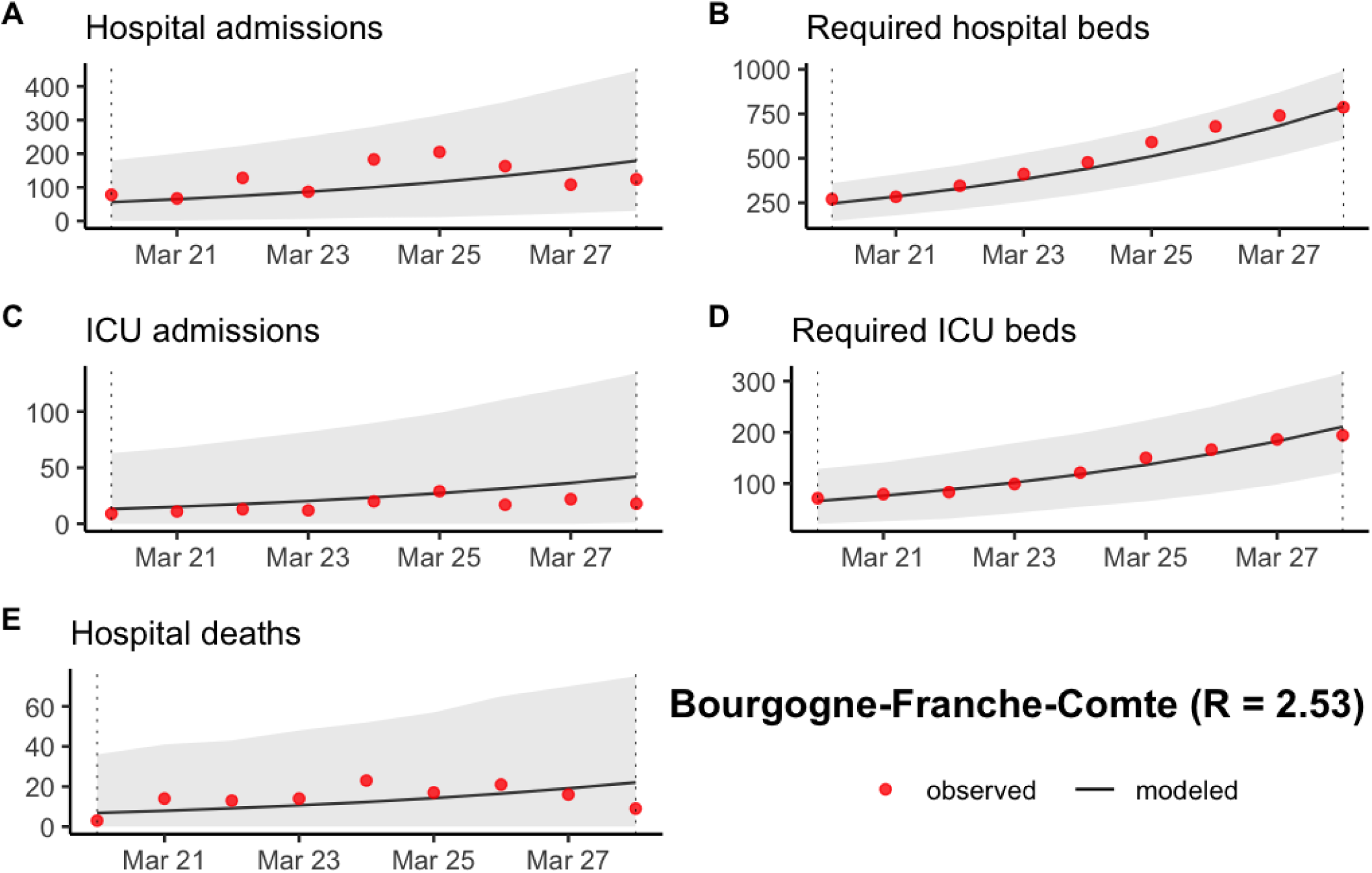
Prediction of (A) the number of new hospitalizations, (B) the number of required hospitalization beds, (C) the number of new ICU admissions, (D) the number of required ICU beds and (E) the number of new hospital deaths in Bourgogne-Franche-Comte between March 20 and March 28 2020. The grey ribbon represents the 95% prediction interval.

**Figure S11:**
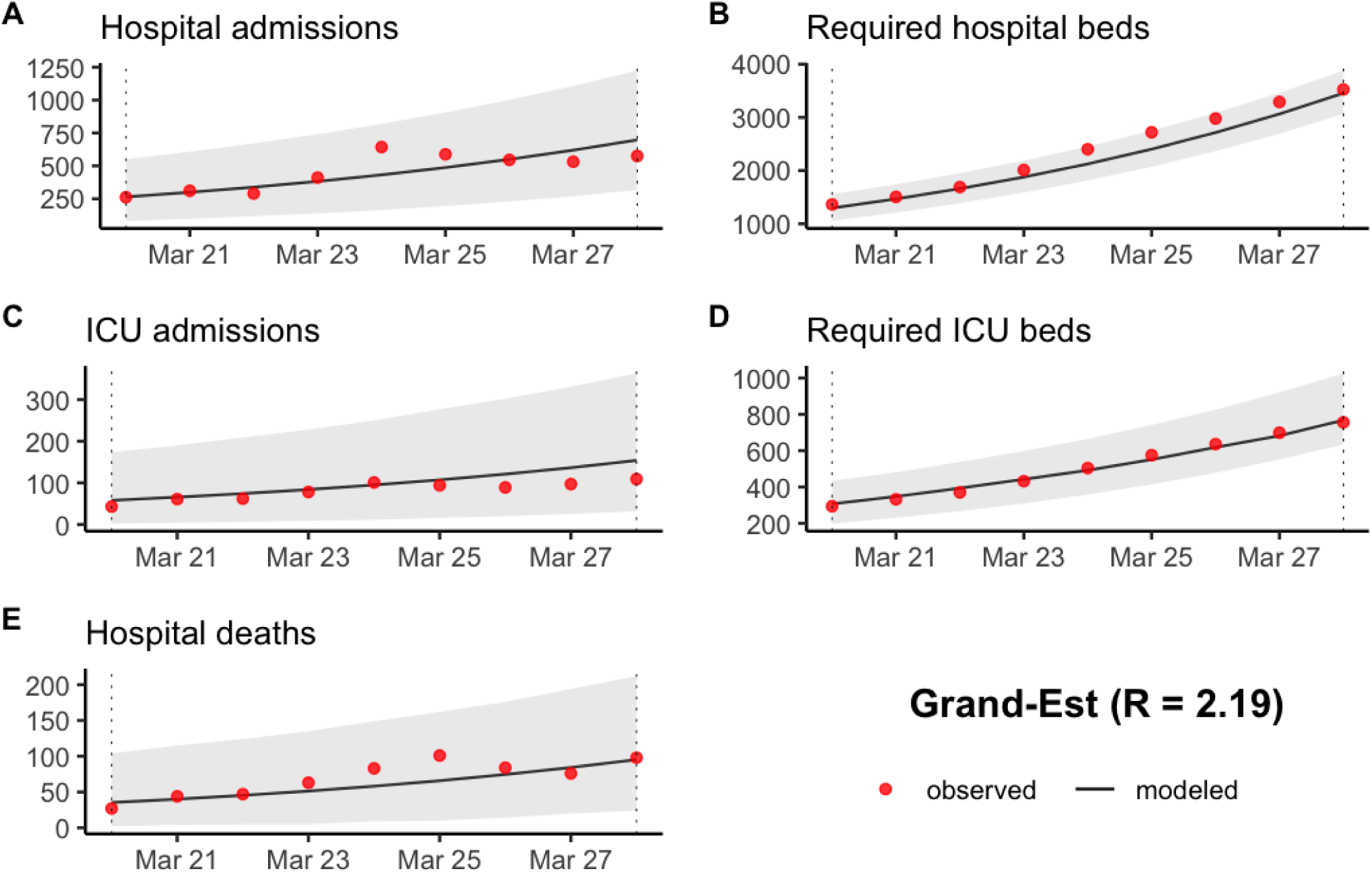
Prediction of (A) the number of new hospitalizations, (B) the number of required hospitalization beds, (C) the number of new ICU admissions, (D) the number of required ICU beds and (E) the number of new hospital deaths in Grand-Est between March 20 and March 28 2020. The grey ribbon represents the 95% prediction interval.

**Figure S12:**
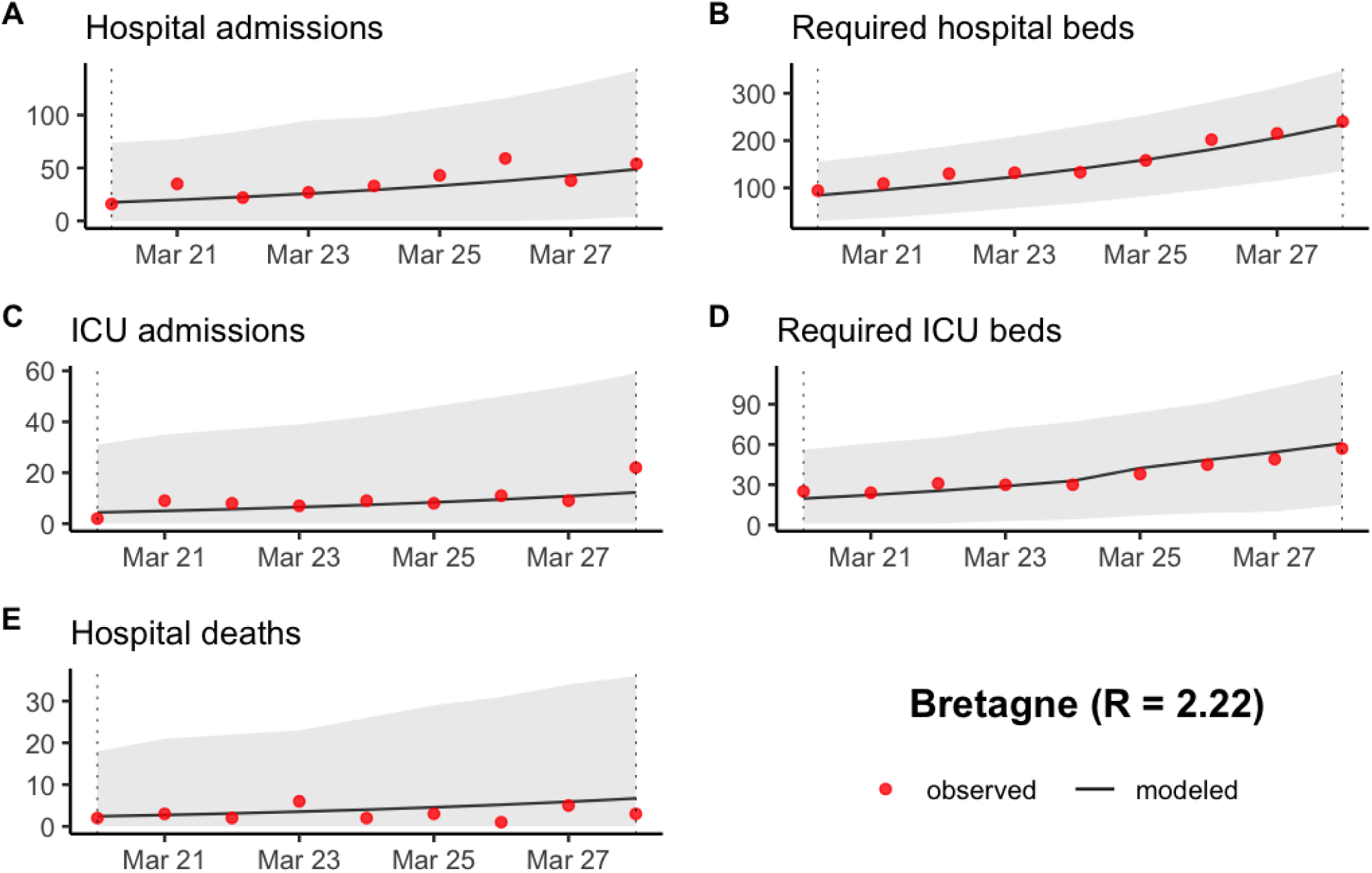
Prediction of (A) the number of new hospitalizations, (B) the number of required hospitalization beds, (C) the number of new ICU admissions, (D) the number of required ICU beds and (E) the number of new hospital deaths in Bretagne between March 20 and March 28 2020. The grey ribbon represents the 95% prediction interval.

**Figure S13:**
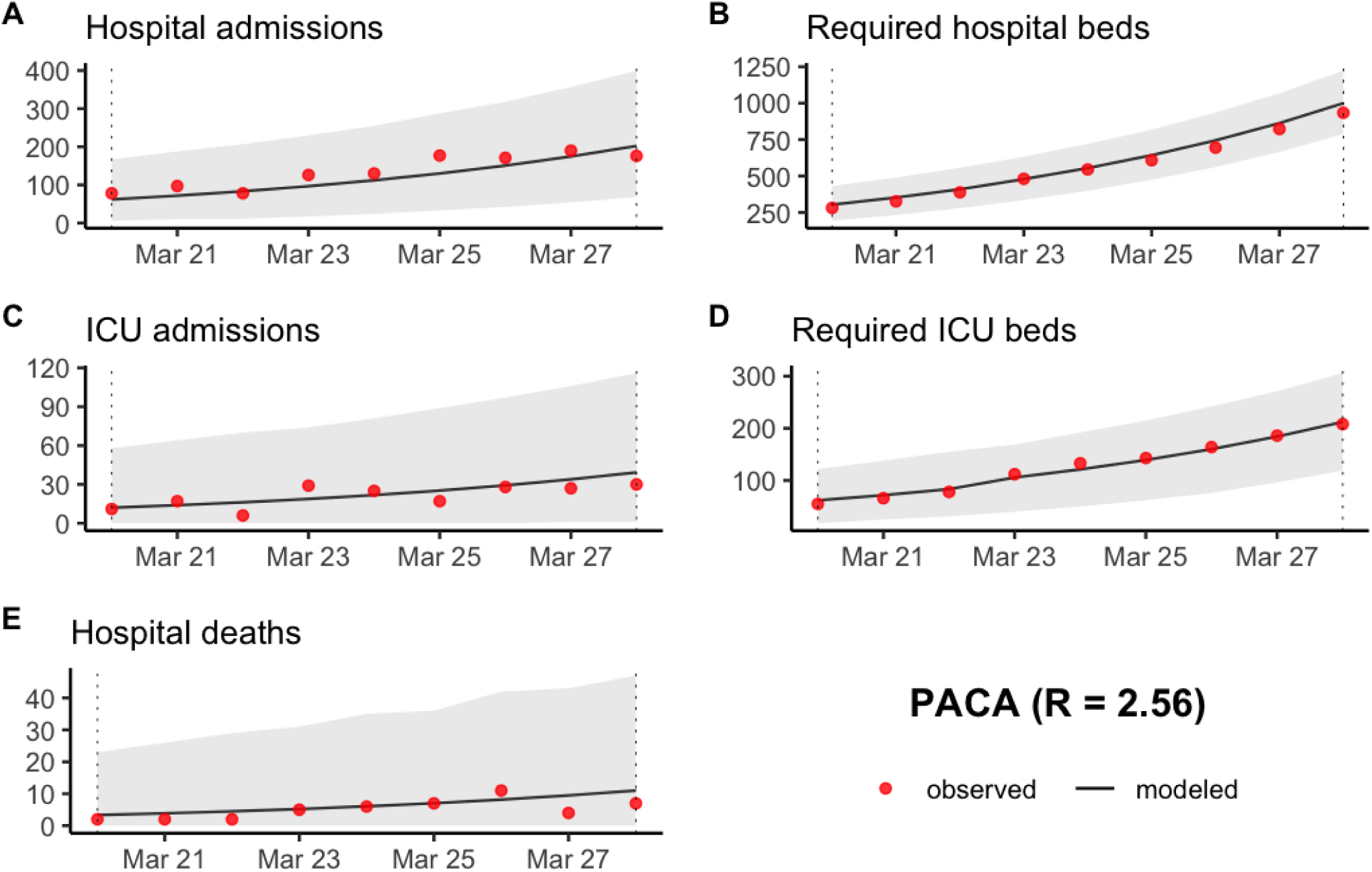
Prediction of (A) the number of new hospitalizations, (B) the number of required hospitalization beds, (C) the number of new ICU admissions, (D) the number of required ICU beds and (E) the number of new hospital deaths in PACA between March 20 and March 28 2020. The grey ribbon represents the 95% prediction interval.

**Figure S14:**
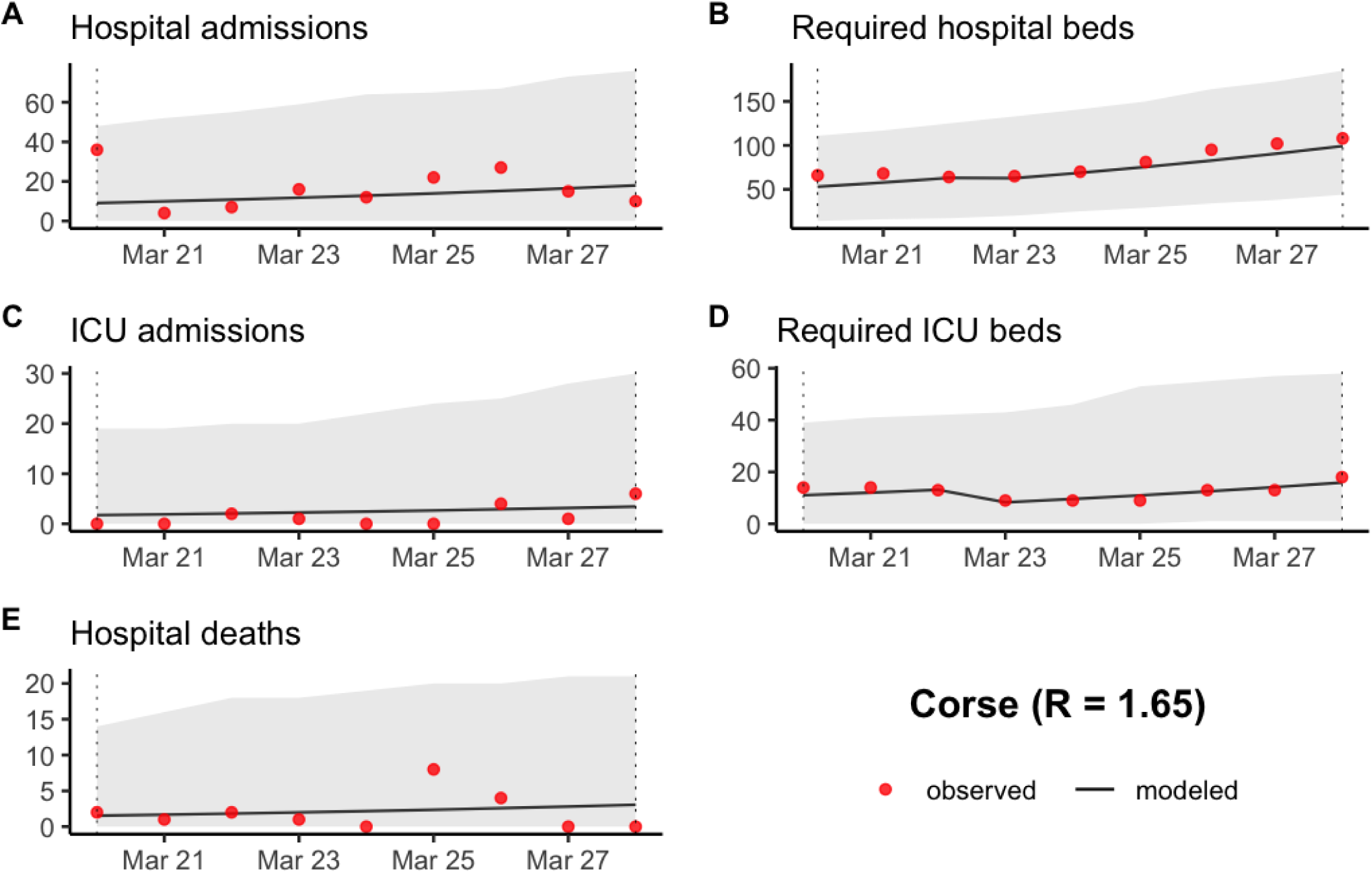
Prediction of (A) the number of new hospitalizations, (B) the number of required hospitalization beds, (C) the number of new ICU admissions, (D) the number of required ICU beds and (E) the number of new hospital deaths in Corse between March 20 and March 28 2020. The grey ribbon represents the 95% prediction interval.

**Figure S15:**
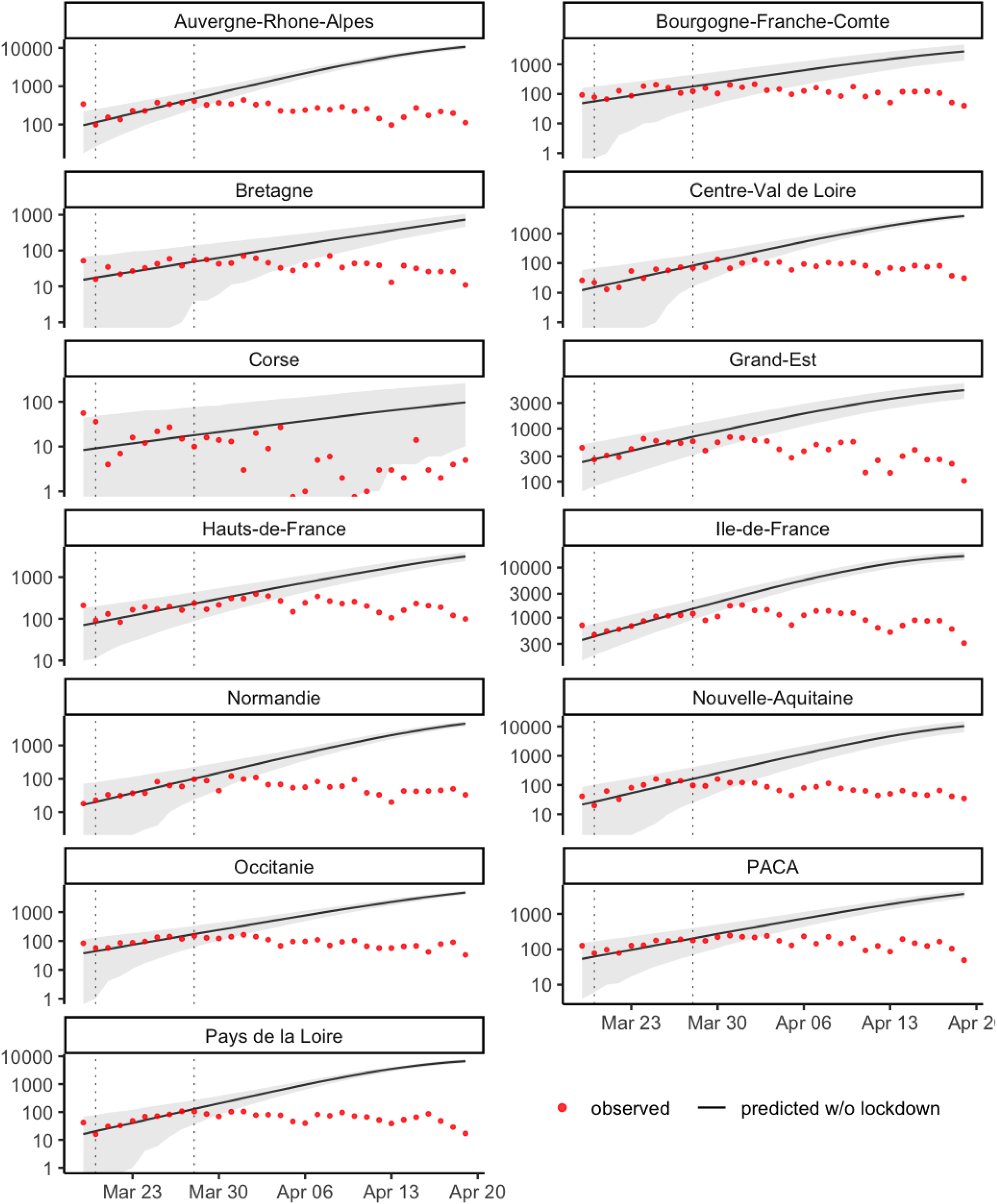
Estimation of the evolution of the number of new hospitalizations by French metropolitan region without lockdown. The grey ribbon represents the 95% prediction interval. The vertical dotted lines indicate the period of time used to estimate the local transmission parameters.

**Figure S16:**
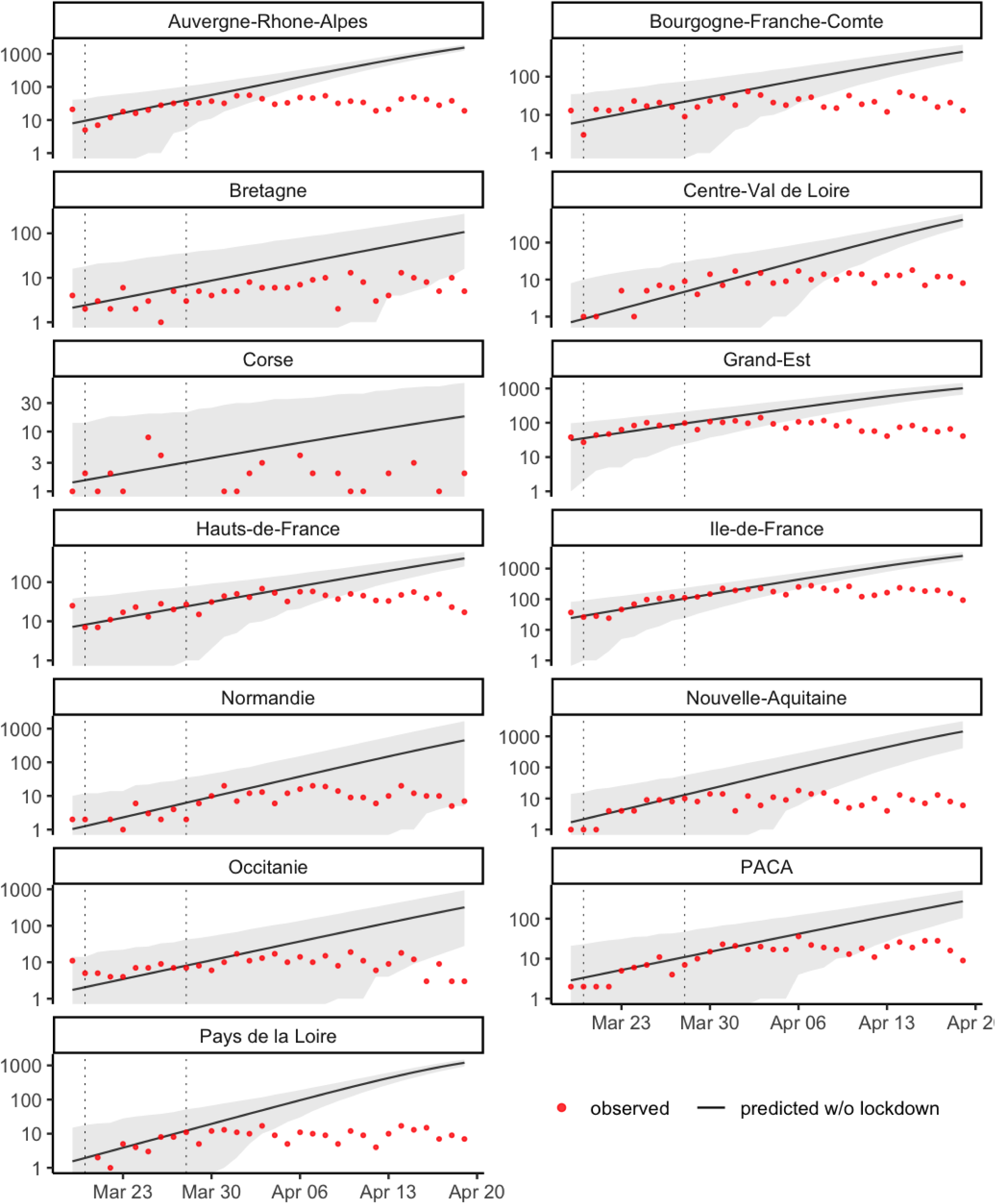
Estimation of the evolution of the number of hospital deaths by French metropolitan region without lockdown. The grey ribbon represents the 95% prediction interval. The vertical dotted lines indicate the period of time used to estimate the local transmission parameters.

